# The longitudinal course of borderline personality disorder: Symptomatology, psychosocial functioning, and factors associated with long-term outcomes

**DOI:** 10.64898/2026.11.20.26368592

**Authors:** Katja I. Seitz, Johannes Zimmermann, Laura Kramer, Paul Hashagen, Corinne Neukel, Christian Schmahl, Katja Bertsch, Sabine C. Herpertz

**Affiliations:** Department of Psychiatry and Psychotherapy, Department of Psychosocial Medicine, Prevention, and Family Health, Medical Faculty, Heidelberg University, Heidelberg, Germany; German Center for Mental Health (DZPG), partner site Mannheim-Heidelberg-Ulm, Germany; Department of Psychology, University of Kassel, Kassel, Germany; Department of Addictive Behaviour and Addiction Medicine, Central Institute of Mental Health, Medical Faculty Mannheim, Heidelberg University, Mannheim, Germany; Department of Psychosomatic Medicine and Psychotherapy, Central Institute of Mental Health, Medical Faculty Mannheim, Heidelberg University, Mannheim, Germany; Department of Psychology I: Clinical Psychology and Psychotherapy, Institute of Psychology, University of Würzburg, Würzburg, Germany; German Center for Mental Health (DZPG), partner site München-Augsburg, Germany

## Abstract

Objective: Longitudinal studies have transformed the understanding of borderline personality disorder (BPD), demonstrating substantial improvement over time. However, seminal findings have largely originated from U.S. cohorts. This study examined the long-term course of BPD and baseline factors associated with clinical and functional outcomes in a large German clinical cohort. Methods: Participants with a lifetime DSM-IV diagnosis of BPD (*N*=410) were assessed at baseline, with follow-up assessments conducted after a mean interval of 8.3 years (*SD*=1.8; range, 5.3–12.0). Changes in categorical diagnosis, dimensional symptom severity, psychosocial functioning, and healthcare utilization were examined using clinician-administered interviews and self-report measures. Baseline factors associated with follow-up outcomes were assessed after adjustment for baseline levels. Results: Follow-up assessments were completed by 186 participants (45.4%); attrition was weakly selective. Among participants with current BPD at baseline, 56.9% no longer met diagnostic criteria according to a 12-month assessment window. Dimensional symptom severity declined substantially across clinician-rated and self-report measures, although individual differences remained partially preserved over time. Psychosocial functioning showed more limited improvement, with 29.5% fulfilling a cross-sectional analogue of recovery including symptomatic remission, employment or education, and a stable partnership. Higher baseline self-reported BPD symptom severity and emotion dysregulation consistently indicated less favorable long-term clinical outcomes, whereas childhood maltreatment, comorbid mental disorders, and further baseline characteristics were not significant predictors. Conclusions: BPD shows substantial long-term improvement, but psychosocial recovery often lags behind symptomatic remission. These findings highlight the importance of treatment approaches that address functioning and quality of life in addition to symptom reduction.

## Introduction

Borderline personality disorder (BPD) is associated with marked functional impairment, psychiatric comorbidity, and high healthcare utilization, contributing to substantial individual and societal burden (1–3). Childhood maltreatment represents an important environmental risk factor in its multifactorial etiology (4).

Longitudinal studies have demonstrated substantial improvements in symptomatology and, to a lesser extent, psychosocial functioning, challenging the longstanding belief that BPD follows an inevitably chronic course (5, 6). Specifically, two landmark studies, the Collaborative Longitudinal Personality Disorders Study (CLPS) and the McLean Study of Adult Development (MSAD), reported high rates of symptomatic remission at 10-year follow-up (85% and 93%, respectively), whereas functional recovery was considerably less common (21% and 50%, respectively) (7–10).

Beyond describing the course of BPD, identifying factors associated with symptomatic remission and psychosocial recovery has important implications for prognosis and treatment planning. However, evidence regarding such factors, including age, gender, childhood maltreatment, and psychiatric comorbidities, is heterogeneous and largely derived from the CLPS and MSAD cohorts (11, 12). Both landmark studies were conducted within the U.S. healthcare system, highlighting the need for independent investigations in other healthcare systems and cultural contexts. Longitudinal studies from non-U.S. settings remain scarce and have generally included fewer than 100 patients with BPD (11, 13), resulting in a limited evidence base in European clinical populations.

To address this gap, the present study characterized the longitudinal course of BPD in a large German clinical cohort, combining nationwide recruitment, multimethod assessment, and comprehensive baseline phenotyping. Using structured clinical interviews and validated self-report measures, we examined longitudinal changes in categorical BPD diagnosis, dimensional symptom severity, psychosocial functioning, and healthcare utilization, as well as baseline factors associated with symptomatic remission and psychosocial recovery. We expected substantial improvements in BPD symptomatology but more modest improvements in psychosocial functioning.

## Methods

### Participants and Procedures

Participants were drawn from a clinical cohort established within the German Research Foundation (DFG)-funded Clinical Research Unit 256, which investigated the neurobiological and psychosocial mechanisms underlying disturbed emotion processing and social interaction in individuals with current or remitted BPD (14). Longitudinal follow-up assessments were conducted as part of a subsequent project (*Symptom Course of BPD: A Follow-up Study*). Data from this follow-up have previously been used to examine the long-term stability of self-reported childhood maltreatment (15).

Between 2012 and 2018, a total of 410 participants with a lifetime diagnosis of BPD were enrolled in the baseline assessment. Participants were recruited throughout Germany through advertisements on social media, newspapers, radio, and podcasts, as well as flyers distributed to general practitioners, psychiatrists, psychotherapists, self-help organizations, and psychiatric inpatient and outpatient services.

Eligibility required a lifetime DSM-IV diagnosis of BPD, defined as the presence of at least five DSM-IV BPD criteria over a period of at least 5 years, as assessed with the International Personality Disorder Examination (IPDE; 16). A current BPD diagnosis at baseline (≥ 5 DSM-IV BPD criteria during the preceding 5 years, including the preceding 12 months) was not required; 65 participants (15.9%) were enrolled with BPD in remission (i.e., below the diagnostic threshold at baseline). Exclusion criteria were current substance use disorder, current psychotic disorder, and current bipolar I disorder, as determined with the Structured Clinical Interview for DSM-IV (SCID-I; 17).

Between 2023 and 2024, follow-up participation was sought among all baseline participants; interviews were conducted primarily via videoconference, occasionally by telephone, by interviewers blinded to baseline status. The baseline and follow-up studies were approved by the Ethics Committee of Heidelberg University and conducted in accordance with the Declaration of Helsinki. All participants provided written informed consent after receiving a complete description of the study; for the 26 participants aged 15-17 years, legal guardian consent was additionally obtained. Participants received compensation for their participation.

### Measures

Unless otherwise specified, assessments were performed at baseline and follow-up using standardized, validated clinician-administered interviews and self-report measures. BPD diagnosis was determined using the IPDE (16). The IPDE yields six BPD outcomes: categorical diagnosis, number of fulfilled DSM-IV BPD criteria, and a dimensional severity score, each assessed for current and lifetime BPD. At baseline, current outcomes reflect symptoms during the preceding 5 years (including the preceding 12 months), whereas lifetime outcomes reflect symptoms present for at least 5 years. At follow-up, outcomes were derived separately for the preceding 12 months and the preceding 5 years, given the established lifetime diagnosis of BPD in all participants. Because the 5-year follow-up window covers a larger proportion of the follow-up period in participants with shorter intervals and thus reaches back closer to baseline, it may attenuate estimates of change; this was examined in sensitivity analyses.

BPD symptom severity was evaluated using complementary clinician-rated and self-report measures, including the Zanarini Rating Scale for Borderline Personality Disorder (ZAN-BPD; 18) and the short version of the Borderline Symptom List (BSL-23; 19). Psychosocial functioning was assessed using standardized questions regarding relationship status, occupational status, and living situation. Healthcare utilization was assessed using standardized questions regarding current psychopharmacological treatment, inpatient psychiatric treatment, and outpatient psychotherapy. Emotion dysregulation, dissociative symptoms, and impulsivity were assessed using the Difficulties in Emotion Regulation Scale (DERS; 20), Dissociative Experiences Scale (DES; 21), and Barratt Impulsiveness Scale,

Version 10 (BIS-10; 22), respectively. Childhood maltreatment was assessed using the Childhood Trauma Questionnaire-Short Form (CTQ-SF; 23). Current and lifetime DSM-IV Axis I disorders were assessed at baseline using the SCID-I (17).

Diagnostic interviews (i.e., IPDE for BPD, ZAN-BPD, and SCID-I) were conducted by trained clinicians. Inter-rater reliability for the IPDE and ZAN-BPD was excellent, with intraclass correlation coefficients (ICCs) of ≥ .98 for the number of BPD criteria and ≥ .91 for BPD symptom severity. All self-report measures demonstrated adequate internal consistency. Detailed descriptions of the instruments, scoring procedures, and psychometric properties are provided in the Supplement.

### Statistical analyses

The analyses examined the longitudinal course of BPD, including diagnosis, symptom severity, psychosocial functioning, and healthcare utilization, as well as baseline demographic and clinical characteristics associated with long-term clinical and functional outcomes. The IPDE-based diagnostic and dimensional indices were the focus of the present analyses; BSL-23 and ZAN-BPD symptom severity provided complementary outcomes, alongside psychosocial functioning and healthcare utilization.

We first evaluated attrition and accounted for missing data (predominantly at follow-up) using multiple imputation (see Supplementary Table S1 for the specification of the imputation models). We then characterized the longitudinal course of BPD by examining changes in diagnosis and symptom severity between baseline and follow-up in terms of both *absolute stability*, defined as mean-level change over time, and *differential (rank-order) stability*, defined as the retention of individual differences over time. We additionally quantified rates of symptomatic remission and recurrence. *Symptomatic remission* was defined as no longer meeting the DSM-IV diagnostic threshold of five criteria at follow-up among participants diagnosed at baseline, and *recurrence* as re-crossing this threshold among participants below it at baseline; both were derived from the baseline-by-follow-up transition tables. We assessed changes in psychosocial functioning, including an approximate, cross-sectional analogue of recovery based on the MSAD concept (9), defined as concurrent symptomatic remission, employment or education, and a stable partnership at follow-up, as well as changes in healthcare utilization. Finally, we examined whether baseline demographic and clinical characteristics were associated with long-term diagnostic, symptomatic, and psychosocial outcomes after adjustment for the respective baseline level. Detailed descriptions of the statistical procedures are provided in the Supplement. All tests were two-tailed with α = .05. Multiple testing was controlled for by using the Benjamini-Hochberg false discovery rate (FDR; 24); all reported *p* values are FDR-adjusted (*p*_FDR_) unless indicated otherwise. Analyses were conducted in R, version 4.4 (25). The analysis script and aggregated outputs are available at https://doi.org/10.5281/zenodo.24310074 (access during peer review: https://zenodo.org/records/24310074?token=eyJhbGciOiJIUzUxMiJ9.eyJpZCI6IjNmZTlhZmUwLWJjYzgtNDdjYy1iNTU4LTIzZmMyMjA4MTc4MyIsImRhdGEiOnt9LCJyYW5kb20iOiIwOTJiZTZiY2VjYjVkYzE4OTY5OWQ0MmViNjc1Yzk4ZSJ9.iGEyhn6zlNhesB5Gs6agfWoc3ldxDAder3tICFhgU5r3c3jvyh_m64B_Ds8HrUSqNER4vlv3MUD19E5d8oQ43g).

## Results

### Follow-Up Sample and Attrition

Follow-up assessments took place *M* = 8.3 years (*SD* = 1.8; range, 5.3–12.0) after baseline. Of the 410 baseline participants, 186 (45.4%) completed the follow-up assessment. Reasons for non-participation were non-response to the initial invitation (37.1%), unavailable or outdated contact information (11.2%), loss to follow-up after initial contact (4.1%), and refusal to participate (2.2%). Baseline sample characteristics are presented in Table 1.

**Table 1.**
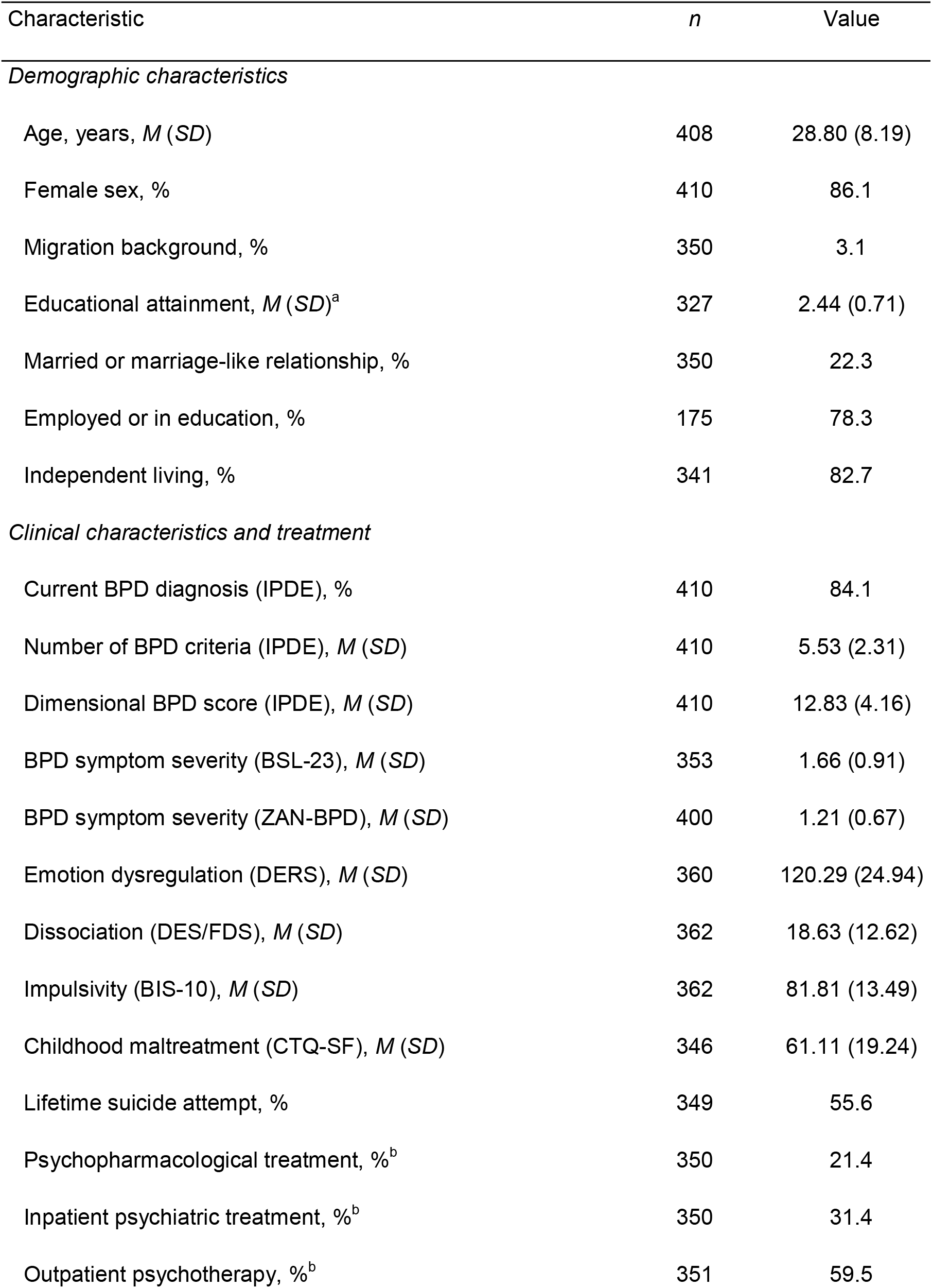
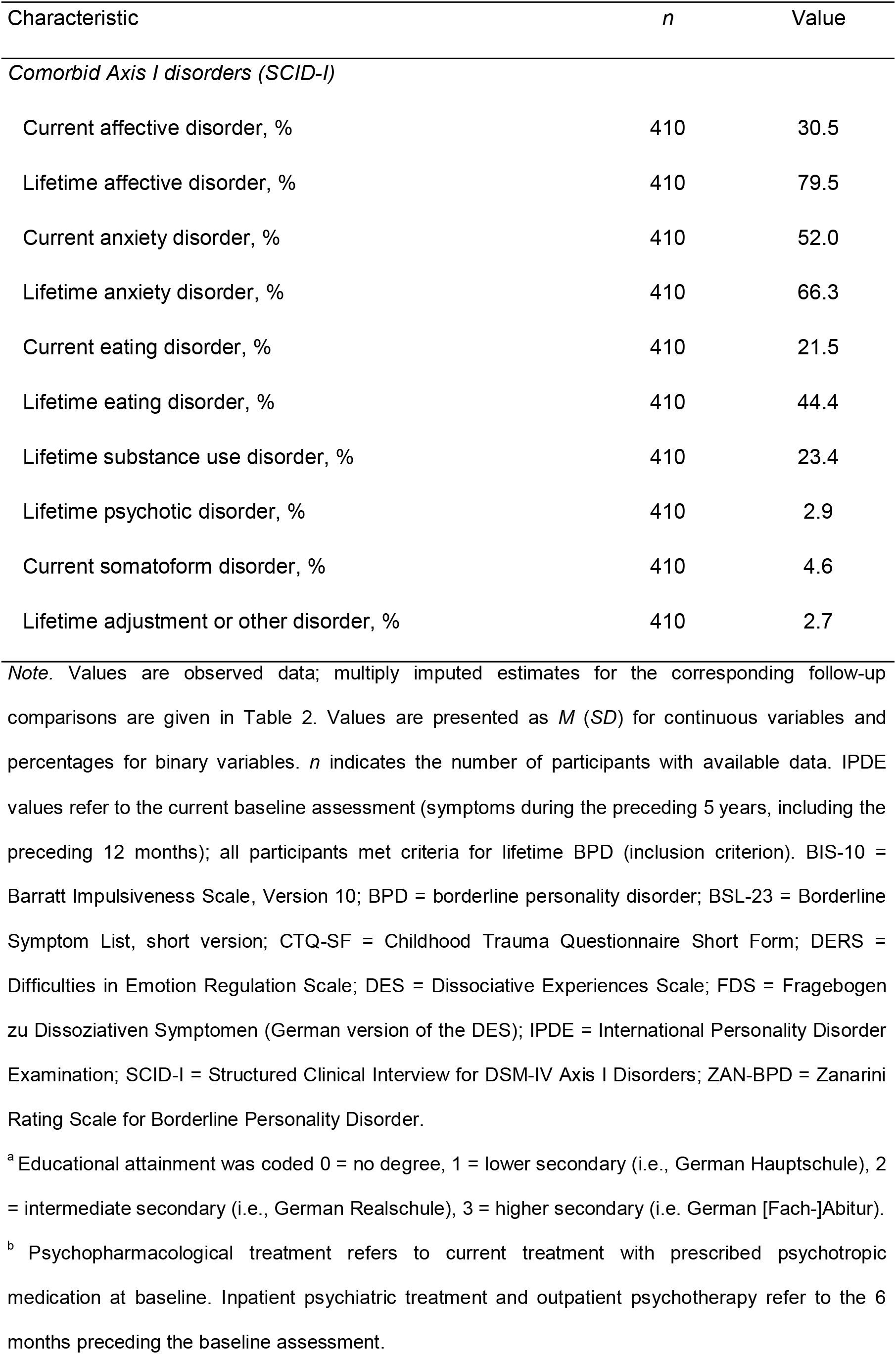
Baseline Sample Characteristics (N = 410)

| Characteristic | *n* | Value |
|---|---|---|
| *Demographic characteristics* |  |  |
| Age, years, *M* (*SD*) | 408 | 28.80 (8.19) |
| Female sex, % | 410 | 86.1 |
| Migration background, % | 350 | 3.1 |
| Educational attainment, *M* (*SD*)<sup>a</sup> | 327 | 2.44 (0.71) |
| Married or marriage-like relationship, % | 350 | 22.3 |
| Employed or in education, % | 175 | 78.3 |
| Independent living, % | 341 | 82.7 |
| *Clinical characteristics and treatment* |  |  |
| Current BPD diagnosis (IPDE), % | 410 | 84.1 |
| Number of BPD criteria (IPDE), *M* (*SD*) | 410 | 5.53 (2.31) |
| Dimensional BPD score (IPDE), *M* (*SD*) | 410 | 12.83 (4.16) |
| BPD symptom severity (BSL-23), *M* (*SD*) | 353 | 1.66 (0.91) |
| BPD symptom severity (ZAN-BPD), *M* (*SD*) | 400 | 1.21 (0.67) |
| Emotion dysregulation (DERS), *M* (*SD*) | 360 | 120.29 (24.94) |
| Dissociation (DES/FDS), *M* (*SD*) | 362 | 18.63 (12.62) |
| Impulsivity (BIS-10), *M* (*SD*) | 362 | 81.81 (13.49) |
| Childhood maltreatment (CTQ-SF), *M* (*SD*) | 346 | 61.11 (19.24) |
| Lifetime suicide attempt, % | 349 | 55.6 |
| Psychopharmacological treatment, %<sup>b</sup> | 350 | 21.4 |
| Inpatient psychiatric treatment, %<sup>b</sup> | 350 | 31.4 |
| Outpatient psychotherapy, %<sup>b</sup> | 351 | 59.5 |
| Characteristic | *n* | Value |
| *Comorbid Axis I disorders (SCID-I)* |  |  |
| Current affective disorder, % | 410 | 30.5 |
| Lifetime affective disorder, % | 410 | 79.5 |
| Current anxiety disorder, % | 410 | 52.0 |
| Lifetime anxiety disorder, % | 410 | 66.3 |
| Current eating disorder, % | 410 | 21.5 |
| Lifetime eating disorder, % | 410 | 44.4 |
| Lifetime substance use disorder, % | 410 | 23.4 |
| Lifetime psychotic disorder, % | 410 | 2.9 |
| Current somatoform disorder, % | 410 | 4.6 |
| Lifetime adjustment or other disorder, % | 410 | 2.7 |

*Note.* Values are observed data; multiply imputed estimates for the corresponding follow-up comparisons are given in Table 2. Values are presented as *M* (*SD*) for continuous variables and percentages for binary variables. *n* indicates the number of participants with available data. IPDE values refer to the current baseline assessment (symptoms during the preceding 5 years, including the preceding 12 months); all participants met criteria for lifetime BPD (inclusion criterion). BIS-10 = Barratt Impulsiveness Scale, Version 10; BPD = borderline personality disorder; BSL-23 = Borderline Symptom List, short version; CTQ-SF = Childhood Trauma Questionnaire Short Form; DERS = Difficulties in Emotion Regulation Scale; DES = Dissociative Experiences Scale; FDS = Fragebogen zu Dissoziativen Symptomen (German version of the DES); IPDE = International Personality Disorder Examination; SCID-I = Structured Clinical Interview for DSM-IV Axis I Disorders; ZAN-BPD = Zanarini Rating Scale for Borderline Personality Disorder.
<sup>a</sup> Educational attainment was coded 0 = no degree, 1 = lower secondary (i.e., German Hauptschule), 2 = intermediate secondary (i.e., German Realschule), 3 = higher secondary (i.e. German [Fach-]Abitur).
<sup>b</sup> Psychopharmacological treatment refers to current treatment with prescribed psychotropic medication at baseline. Inpatient psychiatric treatment and outpatient psychotherapy refer to the 6 months preceding the baseline assessment.

Attrition was only weakly selective: no baseline variable was strongly associated with non-participation (all |*r*| ≤ .16, *p*_FDR_ ≥ .053; Supplementary Table S2). Unless noted otherwise, the results below are based on all 410 participants using multiple imputation; imputation quality checks are reported in Supplementary Table S3.

### Stability of BPD diagnosis and symptoms

Longitudinal changes in BPD diagnostic status and symptom severity are summarized in Table 2 and Figure 1. At baseline, 84.1% of participants met the diagnostic threshold of five or more current BPD criteria. At follow-up, 38.2% met the threshold according to the 12-month window and 58.8% according to the 5-year window. Among participants diagnosed at baseline, 56.9% were in symptomatic remission according to the 12-month window and 35.2% according to the 5-year window; conversely, recurrence occurred in 12.6% (12-month) and 27.3% (5-year) of participants below the diagnostic threshold at baseline. Diagnostic stability was low, with κ = .14 for the 12-month window and κ = .23 for the 5-year window.

**Figure 1.**
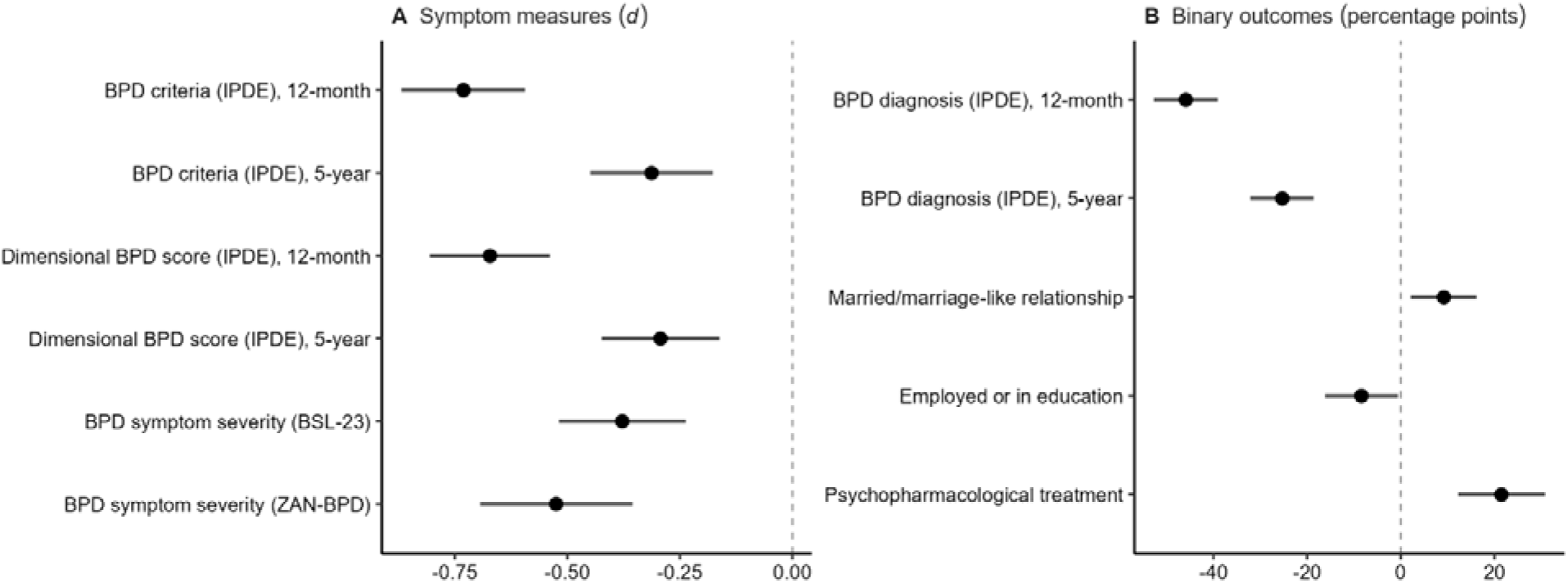
Changes in BPD Symptoms, Diagnostic Status, Psychosocial Functioning, and Healthcare Utilization From Baseline to Follow-Up. *Note*. Points represent pooled estimates of change from baseline to follow-up, with horizontal lines indicating 95% CIs. Panel A shows standardized mean changes (Cohen’s *d*) for continuous symptom measures; negative values indicate lower symptom levels at follow-up. Panel B shows changes in prevalence, expressed as percentage-point differences (follow-up minus baseline), for binary outcomes; negative values indicate lower and positive values higher prevalence at follow-up. Estimates were pooled across 100 multiply imputed data sets. BPD = borderline personality disorder; BSL-23 = Borderline Symptom List, short version; IPDE = International Personality Disorder Examination; ZAN-BPD = Zanarini Rating Scale for Borderline Personality Disorder. Alternative Text for Figure 1: Forest plots display pooled changes in clinical and functional outcomes, with 95% confidence intervals.

**Table 2.** Course of BPD Symptoms, Diagnostic Status, Psychosocial Functioning, and Healthcare Utilization From Baseline to Follow-Up (N = 410, Multiple Imputation)

| Outcome | Baseline | Follow-up | Δ [95% CI] | *p*<sub>FDR</sub> | *d* [95% CI] | *r*/κ [95% CI] |
|---|---|---|---|---|---|---|
| *Diagnostic status* |  |  |  |  |  |  |
| BPD diagnosis (IPDE), 12-month window<sup>a</sup> | 84.1 | 38.2 | -45.9 [-52.7, -39.1] | <.001 | — | .14 [.08, .20] |
| BPD diagnosis (IPDE), 5-year window<sup>a</sup> | 84.1 | 58.8 | -25.3 [-32.1, -18.6] | <.001 | — | .23 [.13, .33] |
| *Symptom measures* |  |  |  |  |  |  |
| Number of BPD criteria (IPDE), 12-month window<sup>a</sup> | 5.53 (2.31) | 3.85 (2.28) | -1.68 [-1.98, -1.37] | <.001 | -.73 [-.87, -.59] | .38 [.28, .48] |
| Number of BPD criteria (IPDE), 5-year window<sup>a</sup> | 5.53 (2.31) | 4.79 (2.42) | -0.74 [-1.06, -0.42] | <.001 | -.31 [-.45, -.18] | .41 [.30, .51] |
| Dimensional BPD score (IPDE), 12-month window<sup>a</sup> | 12.83 (4.16) | 9.98 (4.31) | -2.85 [-3.42, -2.28] | <.001 | -.67 [-.81, -.54] | .40 [.29, .50] |
| Dimensional BPD score (IPDE), 5-year window<sup>a</sup> | 12.83 (4.16) | 11.57 (4.39) | -1.25 [-1.82, -0.68] | <.001 | -.29 [-.42, -.16] | .43 [.32, .53] |
| BPD symptom severity (BSL-23) | 1.65 (0.90) | 1.31 (0.92) | -0.34 [-0.47, -0.22] | <.001 | -.38 [-.52, -.24] | .54 [.44, .63] |
| BPD symptom severity (ZAN-BPD) | 1.22 (0.67) | 0.86 (0.71) | -0.36 [-0.47, -0.25] | <.001 | -.53 [-.69, -.36] | .30 [.17, .42] |
| *Psychosocial functioning* |  |  |  |  |  |  |
| Married or marriage-like relationship | 22.3 | 31.4 | 9.2 [2.1, 16.2] | .013 | — | .36 [.21, .51] |
| Employed or in education | 76.9 | 68.4 | -8.4 [-16.2, -0.6] | .036 | — | .32 [.09, .56] |
| *Healthcare utilization* |  |  |  |  |  |  |
| Psychopharmacological treatment | 21.2 | 42.7 | 21.5 [12.3, 30.8] | <.001 | — | .23 [.07, .39] |
| Inpatient psychiatric treatment<sup>b</sup> | 31.4 | 2.2 | – | – | — | – |
| Outpatient psychotherapy<sup>b</sup> | 59.4 | 40.8 | – | – | — | – |

*Note.* Estimates were pooled across 100 multiply imputed data sets using Rubin's rules. Baseline and follow-up values are *M* (*SD*) for continuous outcomes and prevalence (%) for binary outcomes. Δ represents mean change in raw units for continuous outcomes and percentage-point change for binary outcomes (follow-up minus baseline). *p<sub>FDR</sub>* = Benjamini–Hochberg-adjusted *p* value; *d* = paired difference standardized by the pooled SD across assessments. Stability was quantified using Pearson‘s *r* for continuous outcomes and Cohen‘s κ for binary outcomes. Diagnostic transitions (12-month window): persistence 43.1%, symptomatic remission 56.9%, recurrence 12.6%; (5-year window): persistence 64.8%, remission 35.2%, recurrence 27.3%. BSL-23 = Borderline Symptom List, short version; IPDE = International Personality Disorder Examination; ZAN-BPD = Zanarini Rating Scale for BPD.
<sup>a</sup> Baseline values refer to the current baseline assessment, reflecting BPD symptoms during the preceding 5 years (including the preceding 12 months). Both follow-up windows are compared against this same baseline assessment. All participants met criteria for lifetime BPD at baseline (inclusion criterion; prevalence 100% by definition).
<sup>b</sup> Inpatient psychiatric treatment and outpatient psychotherapy were assessed over different reference periods at baseline (past 6 months) and follow-up (current treatment) and are therefore reported descriptively only.

Mean-level symptom severity declined substantially on all measures (Figure 1A). The number of BPD criteria decreased from *M* = 5.53 (*SD* = 2.31) at baseline to *M* = 3.85 (*SD* = 2.28) in the 12-month window, pooled difference = −1.68, *p*_FDR_ < .001, *d* = −0.73, and to *M* = 4.79 (*SD* = 2.42) in the 5-year window, pooled difference = −0.74, *p*_FDR_ < .001, *d* = −0.31.

The dimensional IPDE score showed the same pattern (12-month: *d* = −0.67; 5-year: *d* = −0.29, both *p*_FDR_ < .001). Self-reported and clinician-rated symptom severity likewise declined, from *M* = 1.65 (*SD* = 0.90) to *M* = 1.31 (*SD* = 0.92) on the BSL-23, *d* = −0.38, *p*_FDR_ < .001, and from *M* = 1.22 (*SD* = 0.67) to *M* = 0.86 (*SD* = 0.71) on the ZAN-BPD, *d* = −0.53, *p*_FDR_ < .001.

Differential stability was moderate for continuous symptom measures: *r* = .38 (12-month window) and *r* = .41 (5-year window) for the number of criteria, *r* = .40 (12-month window) and *r* = .43 (5-year window) for the dimensional score, *r* = .54 for the BSL-23, and *r* = .30 for the ZAN-BPD. Thus, although symptom levels declined markedly, individual differences in symptom severity were partially preserved over the follow-up period.

Sensitivity analyses supported the robustness of the findings (see Supplementary Results and Tables S4-S5).

### Changes in psychosocial functioning and healthcare utilization

Changes in psychosocial functioning were more modest and less uniform than symptomatic improvement (Table 2; Figure 1B). The proportion of participants who were married or in a marriage-like relationship increased from 22.3% to 31.4%, difference = 9.2 percentage points, *p*_FDR_ = .013. Separately, 58.5% of participants reported being in a stable partnership at follow-up, a measure not assessed at baseline that was used for the recovery analogue below. The proportion working or in education decreased from 76.9% to 68.4%, difference = −8.4 percentage points, *p*_FDR_ = .036. This decrease was attributable to the age-related decline in educational enrolment (55 participants were in education at baseline vs. 11 at follow-up); when educational enrolment was excluded, the observed proportion in paid work rose from 46.9% to 62.3%. Independent living was too prevalent at follow-up to be modeled: 96.7% of assessed participants (177/183) lived independently at follow-up, compared with 82.7% at baseline (observed data; denominators vary due to item-level missingness). Following the recovery concept of the MSAD, 29.5% of participants were simultaneously below the diagnostic threshold (12-month window), employed or in education, and in a stable partnership at follow-up (observed data: 29.9%, 55/184). Using the 5-year remission window, the corresponding estimate was 21.0%.

With regard to healthcare utilization, the proportion receiving psychopharmacological treatment increased from 21.2% to 42.7%, difference = 21.5 percentage points, *p*_FDR_ < .001. Inpatient psychiatric treatment and outpatient psychotherapy were assessed over different reference periods at baseline (past 6 months) and follow-up (current treatment) and are therefore reported descriptively: 31.4% versus 2.2% (4/183) for inpatient psychiatric treatment, and 59.4% versus 40.8% for outpatient psychotherapy.

### Predictors of persistence and remission of BPD diagnosis, symptoms, and functional outcomes

Outcomes comprised six IPDE-based measures of BPD diagnostic status and severity (diagnosis, criterion count, and dimensional severity, each for the 12-month and 5-year assessment windows), BPD symptom severity assessed with the BSL-23 and ZAN-BPD, relationship status, employment or education, psychopharmacological treatment, and outpatient psychotherapy. Across all bivariate models adjusting for the baseline level of the respective outcome, 15 of 348 associations (29 baseline predictors × 12 outcomes) remained significant after FDR adjustment within outcome (Table 3; full results in Supplementary Table S6). A consistent pattern emerged: higher baseline self-reported symptom severity (BSL-23) and emotion dysregulation (DERS) predicted a less favorable course on *all six* IPDE outcomes. For the persistence of the diagnosis, each standard deviation on the baseline BSL-23 approximately doubled the odds of meeting diagnostic criteria at follow-up, OR per *SD* = 2.33, *p*_FDR_ < .001 (12-month window), and OR per *SD* = 2.23, *p*_FDR_ < .001 (5-year window). The corresponding estimates for the DERS were OR per *SD* = 2.34, and OR per *SD* = 2.25, both *p*_FDR_ < .001. For the number of criteria and the dimensional score, standardized regression coefficients ranged from β = .24 to β = .34 (all *p*_FDR_ ≤ .032). Higher baseline BSL-23 scores additionally predicted higher follow-up scores on the ZAN-BPD, β = .29, *p*_FDR_ = .032.

**Table 3.**
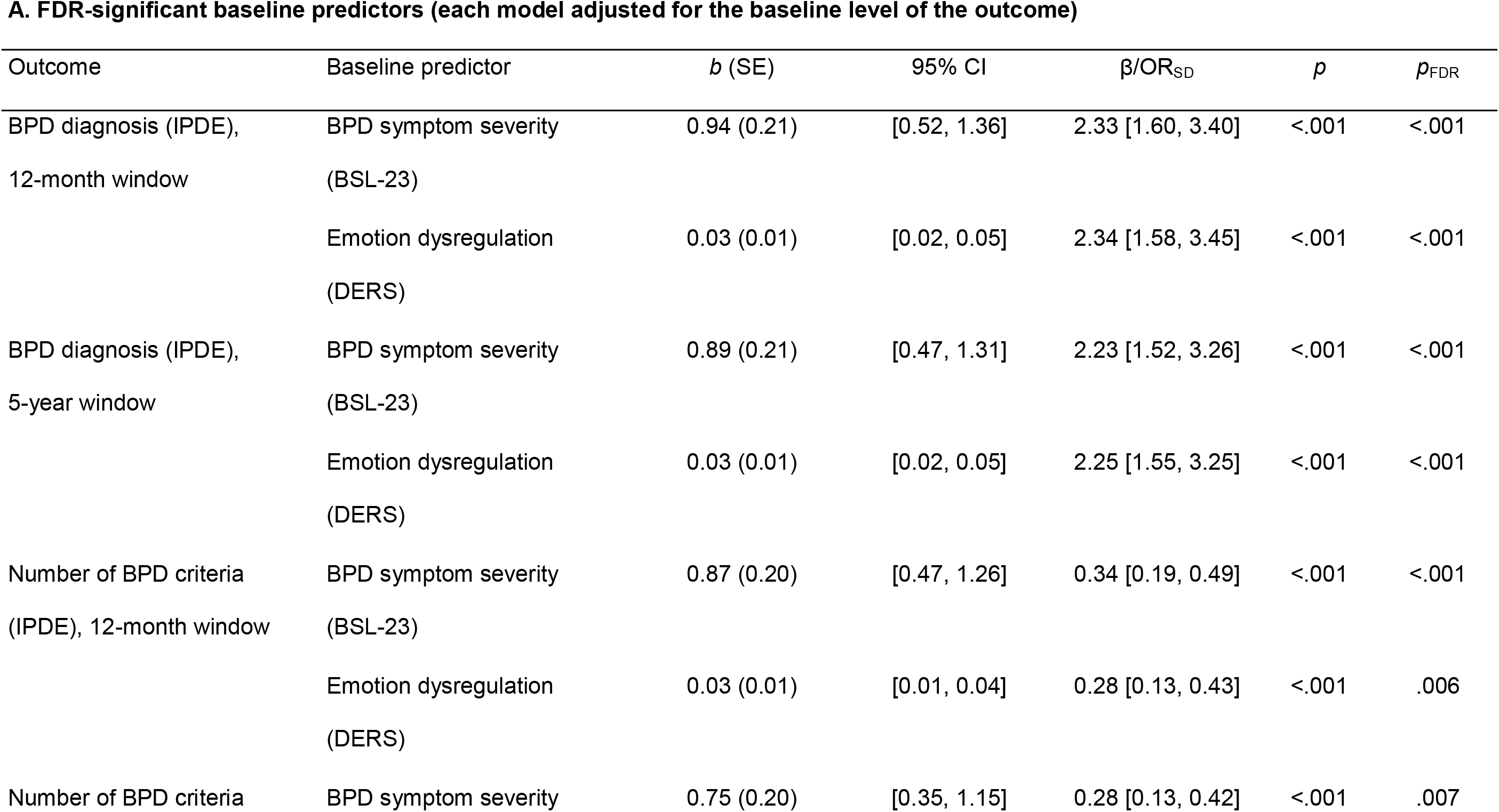
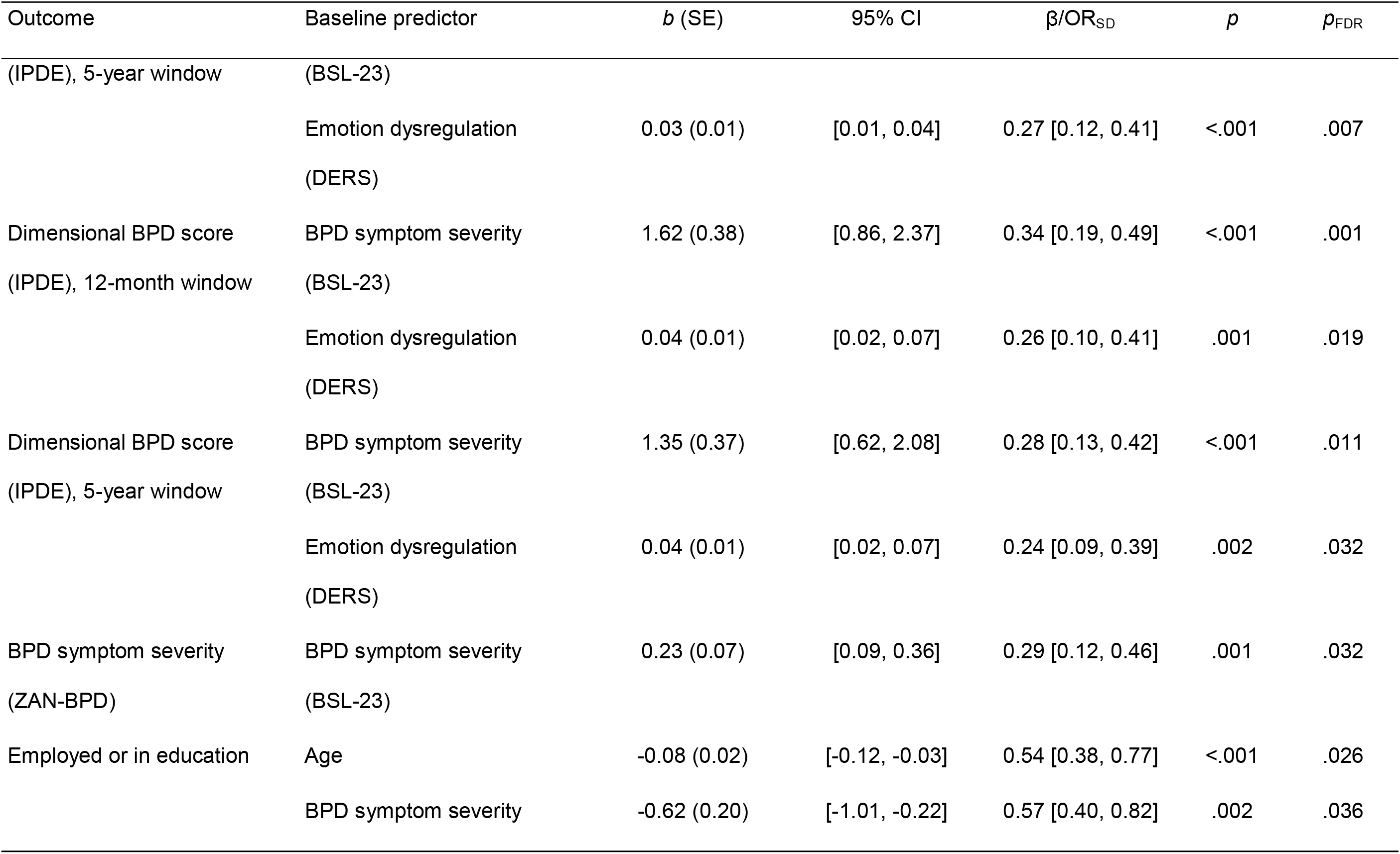
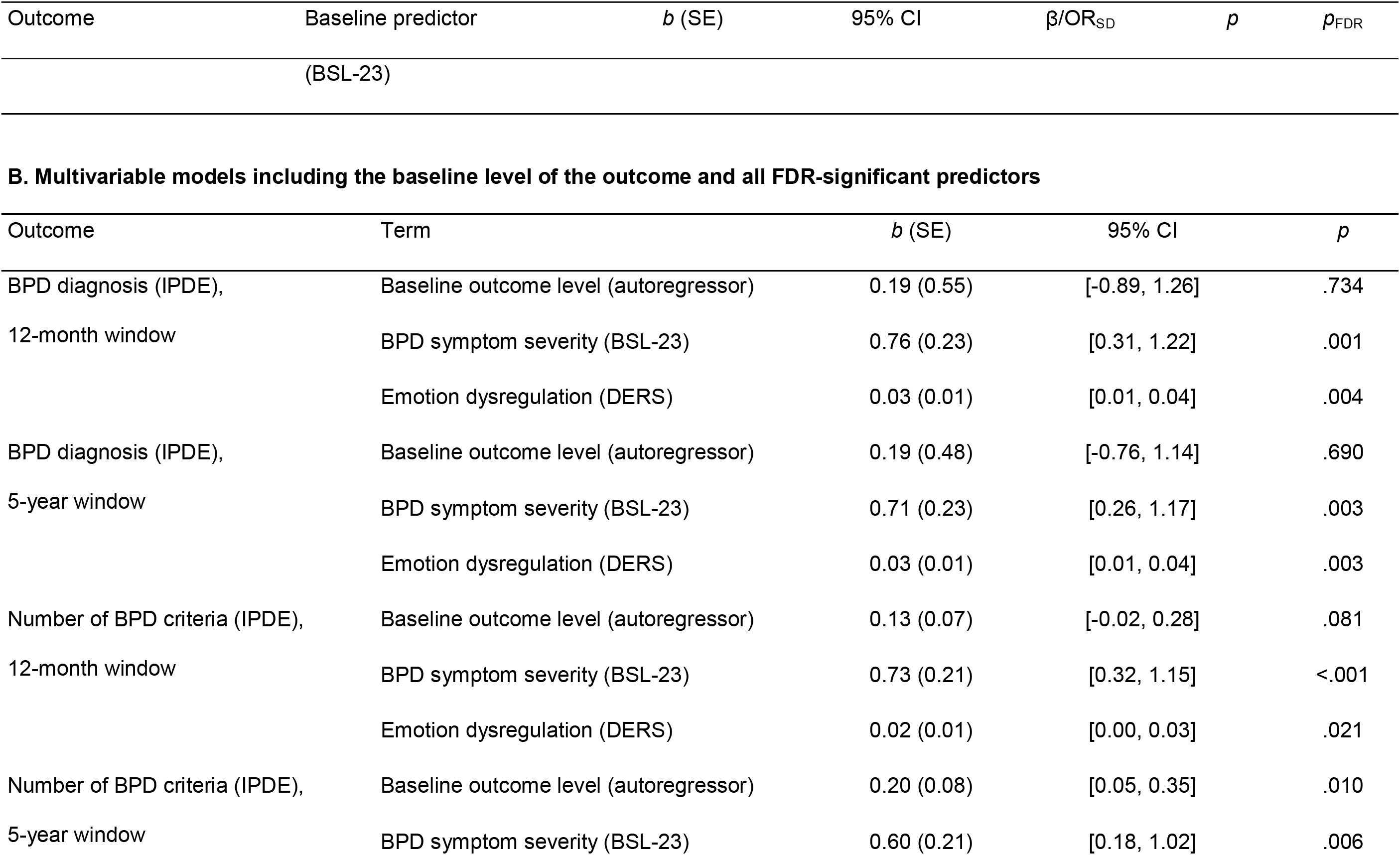
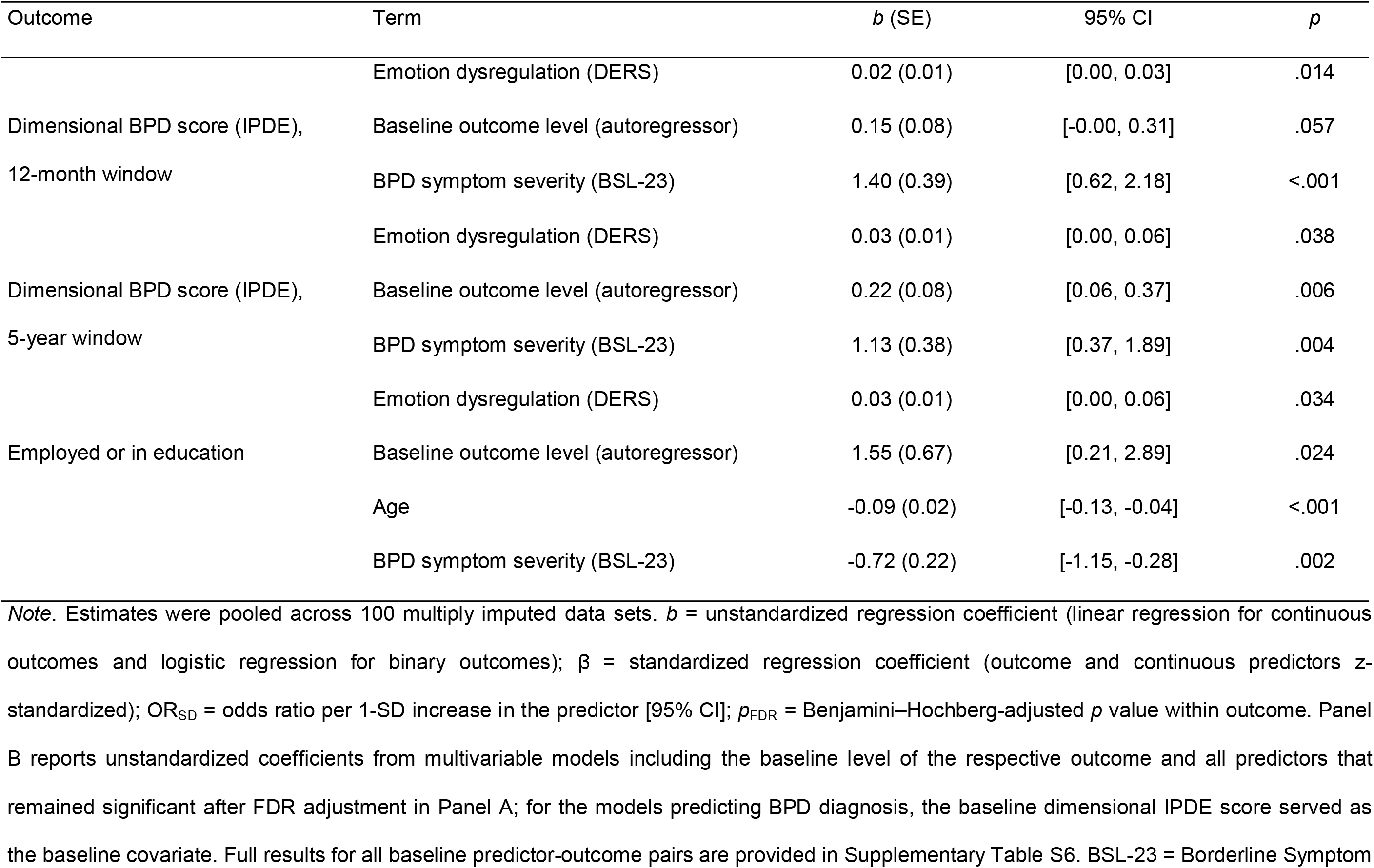
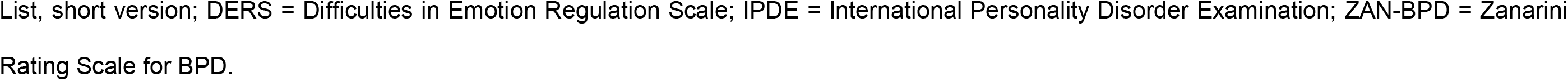
Baseline Predictors of Outcomes at Follow-Up.

| **A. FDR-significant baseline predictors (each model adjusted for the baseline level of the outcome)** |  |  |  |  |  |  |
|---|---|---|---|---|---|---|
| Outcome | Baseline predictor | *b* (SE) | 95% CI | β/OR<sub>SD</sub> | *p* | *p*<sub>FDR</sub> |
| BPD diagnosis (IPDE), 12-month window | BPD symptom severity (BSL-23) | 0.94 (0.21) | [0.52, 1.36] | 2.33 [1.60, 3.40] | <.001 | <.001 |
|  | Emotion dysregulation (DERS) | 0.03 (0.01) | [0.02, 0.05] | 2.34 [1.58, 3.45] | <.001 | <.001 |
| BPD diagnosis (IPDE), 5-year window | BPD symptom severity (BSL-23) | 0.89 (0.21) | [0.47, 1.31] | 2.23 [1.52, 3.26] | <.001 | <.001 |
|  | Emotion dysregulation (DERS) | 0.03 (0.01) | [0.02, 0.05] | 2.25 [1.55, 3.25] | <.001 | <.001 |
| Number of BPD criteria (IPDE), 12-month window | BPD symptom severity (BSL-23) | 0.87 (0.20) | [0.47, 1.26] | 0.34 [0.19, 0.49] | <.001 | <.001 |
|  | Emotion dysregulation (DERS) | 0.03 (0.01) | [0.01, 0.04] | 0.28 [0.13, 0.43] | <.001 | .006 |
| Number of BPD criteria (IPDE), 5-year window | BPD symptom severity (BSL-23) | 0.75 (0.20) | [0.35, 1.15] | 0.28 [0.13, 0.42] | <.001 | .007 |
|  | Emotion dysregulation (DERS) | 0.03 (0.01) | [0.01, 0.04] | 0.27 [0.12, 0.41] | <.001 | .007 |
| Dimensional BPD score (IPDE), 12-month window | BPD symptom severity (BSL-23) | 1.62 (0.38) | [0.86, 2.37] | 0.34 [0.19, 0.49] | <.001 | .001 |
|  | Emotion dysregulation (DERS) | 0.04 (0.01) | [0.02, 0.07] | 0.26 [0.10, 0.41] | .001 | .019 |
| Dimensional BPD score (IPDE), 5-year window | BPD symptom severity (BSL-23) | 1.35 (0.37) | [0.62, 2.08] | 0.28 [0.13, 0.42] | <.001 | .011 |
|  | Emotion dysregulation (DERS) | 0.04 (0.01) | [0.02, 0.07] | 0.24 [0.09, 0.39] | .002 | .032 |
| BPD symptom severity (ZAN-BPD) | BPD symptom severity (BSL-23) | 0.23 (0.07) | [0.09, 0.36] | 0.29 [0.12, 0.46] | .001 | .032 |
| Employed or in education | Age | -0.08 (0.02) | [-0.12, -0.03] | 0.54 [0.38, 0.77] | <.001 | .026 |
|  | BPD symptom severity (BSL-23) | -0.62 (0.20) | [-1.01, -0.22] | 0.57 [0.40, 0.82] | .002 | .036 |
| **B. Multivariable models including the baseline level of the outcome and all FDR-significant predictors** |  |  |  |  |  |  |
| Outcome | Term | *b* (SE) | 95% CI | *p* |  |  |
| BPD diagnosis (IPDE), 12-month window | Baseline outcome level (autoregressor) | 0.19 (0.55) | [-0.89, 1.26] | .734 |  |  |
|  | BPD symptom severity (BSL-23) | 0.76 (0.23) | [0.31, 1.22] | .001 |  |  |
|  | Emotion dysregulation (DERS) | 0.03 (0.01) | [0.01, 0.04] | .004 |  |  |
| BPD diagnosis (IPDE), 5-year window | Baseline outcome level (autoregressor) | 0.19 (0.48) | [-0.76, 1.14] | .690 |  |  |
|  | BPD symptom severity (BSL-23) | 0.71 (0.23) | [0.26, 1.17] | .003 |  |  |
|  | Emotion dysregulation (DERS) | 0.03 (0.01) | [0.01, 0.04] | .003 |  |  |
| Number of BPD criteria (IPDE), 12-month window | Baseline outcome level (autoregressor) | 0.13 (0.07) | [-0.02, 0.28] | .081 |  |  |
|  | BPD symptom severity (BSL-23) | 0.73 (0.21) | [0.32, 1.15] | <.001 |  |  |
|  | Emotion dysregulation (DERS) | 0.02 (0.01) | [0.00, 0.03] | .021 |  |  |
| Number of BPD criteria (IPDE), 5-year window | Baseline outcome level (autoregressor) | 0.20 (0.08) | [0.05, 0.35] | .010 |  |  |
|  | BPD symptom severity (BSL-23) | 0.60 (0.21) | [0.18, 1.02] | .006 |  |  |
|  | Emotion dysregulation (DERS) | 0.02 (0.01) | [0.00, 0.03] | .014 |  |  |
| Dimensional BPD score (IPDE), 12-month window | Baseline outcome level (autoregressor) | 0.15 (0.08) | [-0.00, 0.31] | .057 |  |  |
|  | BPD symptom severity (BSL-23) | 1.40 (0.39) | [0.62, 2.18] | <.001 |  |  |
|  | Emotion dysregulation (DERS) | 0.03 (0.01) | [0.00, 0.06] | .038 |  |  |
| Dimensional BPD score (IPDE), 5-year window | Baseline outcome level (autoregressor) | 0.22 (0.08) | [0.06, 0.37] | .006 |  |  |
|  | BPD symptom severity (BSL-23) | 1.13 (0.38) | [0.37, 1.89] | .004 |  |  |
|  | Emotion dysregulation (DERS) | 0.03 (0.01) | [0.00, 0.06] | .034 |  |  |
| Employed or in education | Baseline outcome level (autoregressor) | 1.55 (0.67) | [0.21, 2.89] | .024 |  |  |
|  | Age | -0.09 (0.02) | [-0.13, -0.04] | <.001 |  |  |
|  | BPD symptom severity (BSL-23) | -0.72 (0.22) | [-1.15, -0.28] | .002 |  |  |

*Note*. Estimates were pooled across 100 multiply imputed data sets. *b* = unstandardized regression coefficient (linear regression for continuous outcomes and logistic regression for binary outcomes); β = standardized regression coefficient (outcome and continuous predictors *z*-standardized); OR<sub>SD</sub> = odds ratio per 1-SD increase in the predictor [95% CI]; *p*<sub>FDR</sub> = Benjamini–Hochberg-adjusted *p* value within outcome. Panel B reports unstandardized coefficients from multivariable models including the baseline level of the respective outcome and all predictors that remained significant after FDR adjustment in Panel A; for the models predicting BPD diagnosis, the baseline dimensional IPDE score served as the baseline covariate. Full results for all baseline predictor-outcome pairs are provided in Supplementary Table S6. BSL-23 = Borderline Symptom List, short version; DERS = Difficulties in Emotion Regulation Scale; IPDE = International Personality Disorder Examination; ZAN-BPD = Zanarini Rating Scale for BPD.

In multivariable models including the autoregressor and both predictors, the BSL-23 and the DERS remained jointly significant for all six IPDE outcomes (e.g., diagnostic status, 12-month window: BSL-23, *b* = 0.76, *p* = .001; DERS, *b* = 0.026, *p* = .004), suggesting non-redundant contributions. Notably, the baseline IPDE level itself was not significantly associated with the outcome in four of these six models once both self-report measures were included (*p* ≥ .057). Younger age and lower baseline BSL-23 scores predicted employment or education at follow-up (Table 3), but not in the complete-case analysis (*n* = 79–84), and should be interpreted cautiously.

Childhood maltreatment (CTQ) was not significantly associated with any outcome after FDR adjustment: its associations with the IPDE outcomes were small and positive, β = .13–.15 (diagnosis: OR per *SD* ≤ 1.44), with unadjusted *p* = .007–.032 and *p*_FDR_ ≥ .063. Complete-case zero-order correlations were of similar magnitude (*r* = .17–.18 with the follow-up criterion count, *n* = 163), indicating that this largely null result is not attributable to the imputation procedure. No other baseline factors showed FDR-significant associations with the outcomes.

## Discussion

The present study extends prior research on the long-term course of BPD by examining one of the largest European cohorts of individuals with BPD over a mean follow-up of 8.3 years. Three principal findings emerged. First, substantial improvements occurred across categorical and dimensional measures of BPD, consistent with previous evidence that BPD is not inevitably chronic. Second, symptomatic remission substantially exceeded psychosocial recovery, underscoring that diagnostic remission represents an important but incomplete marker of recovery. Third, baseline self-reported BPD symptom severity and emotion dysregulation consistently predicted long-term clinical outcomes, whereas childhood maltreatment and most other baseline characteristics showed no significant associations after correction for multiple testing. Together, these findings extend previous longitudinal research and highlight important methodological considerations in assessing the course of BPD.

Our findings support the favorable long-term course of BPD while demonstrating the importance of how remission is defined and assessed. The 57% remission rate approximately 8 years after baseline is lower than the cumulative rates of 85% in the CLPS (7) and 93% in the MSAD (9); critically, however, these findings reflect fundamentally different approaches to characterizing longitudinal remission. Repeated assessments in the CLPS and MSAD identified individuals who achieved remission at any point during the 10-year follow-up, whereas our single follow-up assessment provides an estimate of remission during the preceding 12 months and cannot account for intervening periods of remission and recurrence. Moreover, our remission threshold (< 5 BPD criteria) was less stringent than that of the CLPS (< 2 criteria) and required a shorter remission period than the MSAD (12 months vs. 2 years). Notably, our estimate closely aligns with the 60% remission rate reported in a recent meta-analysis of prospective longitudinal studies across nine countries, which, like our study, assessed remission at a specific follow-up point after 5 to 15 years rather than cumulatively across repeated assessments (11). Beyond differences in remission definitions, remission rates may also vary as a function of cohort composition. Unlike the CLPS and MSAD, which recruited treatment-seeking patients from clinical settings, our nationwide cohort was not restricted to patients receiving specialized care and may capture a broader spectrum of the long-term course of BPD within the German healthcare system.

Consistent with the CLPS (7) and MSAD (9), symptomatic remission was considerably more pronounced than improvement in psychosocial functioning. Approximately 30% of participants fulfilled our cross-sectional analogue of recovery at follow-up, despite a relatively lenient vocational criterion including both employment and education. Although not directly comparable with cumulative recovery estimates from MSAD, this finding supports the conclusion that improvement in core symptoms does not necessarily coincide with recovery in psychosocial functioning. Persistent psychosocial difficulties after symptomatic remission may reflect not only residual impairments but also the consequences of a prolonged illness trajectory, including disrupted educational and occupational development, strained interpersonal relationships, and stigma (26–28) that may shape social and occupational opportunities even after symptoms have substantially improved. Despite substantial symptomatic improvement, psychopharmacological treatment increased over follow-up, potentially reflecting persistent psychiatric comorbidities or changing treatment needs with age, although pharmacotherapy is not recommended for the core symptoms of BPD. Clinically, these findings emphasize that symptom reduction alone should not be regarded as the sole treatment goal; interventions targeting relationships, occupational participation, and broader social functioning remain essential even after patients no longer meet diagnostic criteria for BPD.

Baseline self-reported BPD symptom severity in the preceding week (BSL-23) and emotion dysregulation (DERS) emerged as the most consistent factors associated with interviewer-assessed categorical BPD diagnosis and dimensional indicators of BPD severity (IPDE) at follow-up. Notably, after accounting for these dimensional self-report measures, interviewer-assessed BPD severity provided limited incremental information in most multivariable models. This finding suggests that dimensional self-report measures capture prognostically relevant variance beyond that captured by the interview-based assessment, possibly because the latter reflected current severity less precisely and was range-restricted by the inclusion criteria. The stronger associations of self-report measures may arise from complementary aspects of assessment, including the greater reliability of multi-item questionnaires, differences in assessment windows, or differential sensitivity to state-dependent and more enduring features of BPD pathology. Given its prognostic relevance, emotion dysregulation may represent a promising treatment target for interventions such as dialectical behavior therapy (DBT). In contrast to some previous findings (29, 30), childhood maltreatment showed only small positive associations with follow-up clinical outcomes that did not survive correction for multiple testing. Although childhood maltreatment has been associated with BPD development and severity (31), its role in the long-term course appears more heterogeneous and may depend on subsequent developmental, interpersonal, and treatment-related factors.

Beyond clinical findings, our study highlights methodological considerations relevant to longitudinal research on BPD. Our window-coverage sensitivity analysis supports the use of the 12-month remission window as the primary approach while illustrating how remission estimates are influenced by temporal definitions. Future studies with repeated assessments should further characterize change at the level of individual BPD criteria, which may follow distinct trajectories, with acute symptoms such as self-harm potentially remitting faster than more temperamental features (32, 33). In addition, the lower stability of categorical diagnoses, which partly reflects the large shift in prevalence that constrains κ, underscores the value of complementing categorical outcomes with dimensional assessments when characterizing the long-term course of BPD.

Several limitations warrant consideration. The single follow-up assessment limits conclusions regarding the timing and continuity of remission and recurrence. Inpatient psychiatric treatment and outpatient psychotherapy were assessed over different reference periods at baseline (past 6 months) and follow-up (current treatment), potentially overstating reductions in treatment utilization; these findings are therefore reported descriptively only. Although nationwide recruitment extended beyond specialized treatment settings, generalizability may be limited by the inclusion of individuals whose qualifying BPD symptoms occurred many years before baseline, the exclusion of common comorbidities such as substance use disorders (34), and the uncertain representativeness of the 40.8% outpatient psychotherapy utilization rate at follow-up in the absence of population-based German data. Employment status was missing for more than half of the sample at baseline, potentially affecting functional outcome estimates alongside attrition; however, attrition was only weakly selective and its potential impact was addressed using multiple imputation and full-information maximum likelihood, both of which assume that non-participation is ignorable given the baseline variables. Finally, our pragmatic recovery analogue may not capture the full complexity of psychosocial functioning or alternative pathways to recovery.

In conclusion, our findings highlight that BPD should not be conceptualized as an inevitably chronic disorder while emphasizing that recovery extends beyond symptomatic remission. A comprehensive assessment of long-term outcomes should integrate symptomatic change, psychosocial functioning, and individual prognostic factors rather than relying on diagnostic status alone. This multidimensional perspective supports treatment approaches that extend beyond symptom reduction to promote meaningful psychosocial recovery and quality of life.

## Data Availability

The individual-level data are not publicly available because participants did not consent to public data sharing; they are available from the corresponding author upon reasonable request.

https://doi.org/10.5281/zenodo.24310074

## Acknowledgments

This work was supported by the German Research Foundation under Grant No.190034061; and the German Center for Mental Health (Deutsches Zentrum für Psychische Gesundheit, DZPG) under Grant No. DZPG 01EE2504B. The data storage service SDS@hd was supported by the Ministry of Science, Research and the Arts Baden-Württemberg and German Research Foundation under Grant No. INST 35/1503-1 FUGG. A large language model (Claude, Anthropic) was used to cross-check the analysis script against the reported results and to suggest language edits; all analyses, results, and text were verified by the authors.

