## Supplement for "The longitudinal course of borderline personality disorder: Symptomatology, psychosocial functioning, and factors associated with long-term outcomes"

**Supplementary Information**

**Supplementary Methods**

*Clinician-Administered Interviews and Self-Report Measures*

Clinician-administered interviews included the International Personality Disorder Examination (IPDE; 1), the Zanarini Rating Scale for Borderline Personality Disorder (ZAN-BPD; 2), and the Structured Clinical Interview for DSM-IV Axis I Disorders (SCID-I; 3). All interviews were conducted by trained clinicians who underwent standardized training and received regular supervision. Inter-rater reliability was established at baseline using randomly selected videotaped diagnostic interviews. According to widely accepted standards (4), inter-rater reliability, quantified as intraclass correlation coefficients (ICCs), was consistently excellent for both the number of BPD criteria assessed with the IPDE (ICC ≥ .98) and BPD symptom severity assessed with the ZAN-BPD (ICC ≥ .91).

Borderline personality disorder (BPD) diagnosis was established using the IPDE (1). The IPDE is a clinician-administered, semi-structured interview assessing personality disorders according to the *International Statistical Classification of Diseases and Related Health Problems, Tenth Revision* (ICD-10; 5) and the *Diagnostic and Statistical Manual of Mental Disorders, Fourth Edition* (DSM-IV; 6). The IPDE permits the assessment of both current and lifetime (past) personality disorders. Current diagnoses require characteristic behaviors or traits to have been present for at least 5 years, including during the preceding 12 months, whereas lifetime diagnoses meet the same diagnostic criteria but do not require the presence of symptoms during the preceding 12 months. BPD is assessed using 15 interview items covering the nine DSM-IV diagnostic criteria. Each item is rated on a three-point scale, including 0 (*absent or within normal limits*), 1 (*subthreshold*), and 2 (*criterion-level*). Item scores ≥ 1 require the corresponding behavior or trait to have been present for at least 5 years. The IPDE provides both categorical and dimensional indices of BPD. Categorically, diagnoses are classified as definite (≥ 5 criteria), probable (4 criteria), or negative (≤ 3 criteria). Dimensionally, the IPDE yields the number of concurrently fulfilled DSM-IV BPD criteria (range: 0-9) and a dimensional severity score calculated as the sum of the nine DSM-IV criterion scores (range: 0-18), with higher scores indicating greater symptom severity.

Severity of BPD symptoms was assessed using the ZAN-BPD (2) and the short version of the Borderline Symptom List (BSL-23; 7). The ZAN-BPD is a clinician-administered interview assessing the severity of the nine DSM-IV BPD symptom domains during the preceding week. Each domain is rated on a 5-point scale ranging from 0 (*no symptoms*) to 4 (*severe symptoms*). The interview yields a total score (range: 0-36; reported here as the mean of the nine items, range: 0-4) and four sector scores representing affective, cognitive, impulsive, and interpersonal symptom severity, with higher scores indicating greater symptom severity. Internal consistency was good in previous studies (Cronbach’s α = .85) (2) and ranged from acceptable to good in the present sample (Cronbach’s α = .78 at baseline and .84 at follow-up; Table S7). The BSL-23 is a 23-item self-report questionnaire assessing BPD symptom severity during the preceding week. Items are rated on a 5-point scale ranging from 0 (*not at all*) to 4 (*very strong*). The total score (range: 0.0-4.0) is calculated as the mean of all valid item scores, with higher scores reflecting greater symptom severity. Internal consistency was excellent in both previous studies (Cronbach’s α = .94-.97) (7) and the present sample (Cronbach’s α = .95 at baseline and .96 at follow-up).

Emotion dysregulation was measured using the German version of the Difficulties in Emotion Regulation Scale (DERS; 8). The DERS is a 36-item self-report questionnaire assessing six domains of emotion dysregulation: nonacceptance of emotional responses, difficulties engaging in goal-directed behavior, impulse control difficulties, lack of emotional awareness, limited access to emotion regulation strategies, and lack of emotional clarity. Items are rated on a 5-point scale ranging from 1 (*almost never*) to 5 (*almost always*). The questionnaire yields a total score (range: 36-180), with higher scores reflecting greater emotion dysregulation. Internal consistency was excellent in both previous studies (Cronbach’s α = .93) (9) and the present sample (Cronbach’s α = .93 at baseline).

Dissociation was assessed using the German adaptation of the Dissociative Experiences Scale (DES; 10), the Fragebogen zu dissoziativen Symptomen (FDS; 11). The FDS is a 44-item self-report questionnaire comprising the original 28 DES items, which assess amnesia, absorption, and depersonalization/derealization, as well as 16 additional items assessing conversion symptoms. Items are rated on a visual analogue scale ranging from 0% (*never*) to 100% (*always*). The dissociation score was calculated as the mean of all 44 FDS items (range: 0-100), with higher scores indicating greater dissociative symptom severity. Internal consistency of the FDS was excellent in both previous studies (Cronbach’s α = .93) (11) and the present sample (Cronbach’s α = .94 at baseline), and was thus comparable to that of the original DES.

Impulsivity was evaluated using the Barratt Impulsiveness Scale, Version 10 (BIS-10; 12; unpublished German translation by S. C. Herpertz, R. Lohmann, and T. Lohmann). The BIS-10 is a 34-item self-report questionnaire assessing trait impulsivity. Items are rated on a 4-point scale ranging from 1 (*rarely/never*) to 4 (*almost always/always*). The questionnaire yields a total score (range: 34-136) and three subscale scores assessing attentional, motor, and non-planning impulsivity, with higher scores indicating greater impulsivity. Internal consistency in the present sample was good (Cronbach’s α = .85 at baseline).

Childhood maltreatment was assessed using the German version of the Childhood Trauma Questionnaire–Short Form (CTQ-SF) (13). The CTQ-SF is a self-report questionnaire comprising five 5-item subscales assessing emotional abuse, physical abuse, sexual abuse, emotional neglect, and physical neglect. Items are rated on a 5-point scale ranging from 1 (*not at all*) to 5 (*very often*). The questionnaire yields a total score (range: 25-125) and five subscale scores (range: 5-25), with higher scores indicating more severe abuse or neglect. Recommended cutoff scores for clinically significant childhood maltreatment are ≥8 for sexual abuse, ≥10 for physical abuse and physical neglect, ≥13 for emotional abuse, and ≥15 for emotional neglect. Internal consistency was good in a recent meta-analysis (Cronbach’s α = .89) (14) and excellent in the present sample (Cronbach’s α = .93 at baseline).

Comorbid mental disorders were determined at baseline using the German version of the SCID-I (15). The SCID-I is a semi-structured, clinician-administered interview for the assessment of current and lifetime DSM-IV Axis I disorders.

*Statistical Analyses*

The primary analyses used multiple imputation by chained equations (m = 100 imputations; mice package; 16), providing valid inference under the missing-at-random assumption, which was made plausible by including attrition-related variables in the imputation models (17). The four IPDE follow-up summary scores – the criterion count and dimensional severity score for each of the 12-month and 5-year assessment windows – were imputed jointly as one block using multivariate predictive mean matching (18) to preserve their internal consistency. Specifically, each non-participant received the complete, internally consistent score vector of an observed donor matched on baseline predictors. Because categorical BPD diagnosis is determined directly from the criterion count (≥ 5 criteria), it was derived from the imputed criterion count and not imputed separately. All other variables were imputed univariately using predictive mean matching. Follow-up variables were imputed from baseline information only. Imputation-model predictors were selected separately for each target variable based on associations with the target's observed values (|*r*| ≥ .20) or its missingness indicator (|*r*| ≥ .10), supplemented by the corresponding baseline score and the follow-up interval. The follow-up interval was reconstructed as a complete design variable from the baseline assessment dates and predicted attrition. Further baseline instruments were screened as candidate auxiliary variables; none met the inclusion threshold.

The IPDE at follow-up provided two assessment windows, covering the past 12 months and the past 5 years (see *Measures* in the main manuscript). Both were modelled against the same baseline assessment, which had established the presence of criteria for at least 5 years including the past 12 months. Because the 5-year window covers a proportion of the follow-up period that depends on the individual follow-up interval (94% at 5.3 years, 42% at 12 years) and thus reaches back closer to baseline for shorter intervals, we tested this window-coverage mechanism directly as a sensitivity analysis (see below). The differing window lengths at baseline and follow-up primarily affect estimates of mean-level change; as the baseline assessment is the stricter one, the observed symptomatic decline is, if anything, underestimated. Full specification of the imputation models, the attrition analysis, and quality checks comparing observed and imputed distributions are provided in Supplementary Tables S1–S3. Estimates were pooled across imputations by Rubin's rules (19), with correlations pooled after Fisher *z* transformation, prevalences pooled on the logit scale, and standard errors for differences of dependent proportions based on the McNemar variance; pooled Wald tests used Barnard–Rubin degrees of freedom.

We characterized the course of BPD in terms of *absolute stability*, that is, mean-level change between baseline and follow-up, and *differential (or rank-order) stability*, that is, the retention of individual differences over time. For continuous outcomes, mean-level change was quantified as the paired difference between occasions and standardized as *d* = difference / pooled *SD* of the two occasions (20); differential stability was quantified as the Pearson correlation between baseline and follow-up scores. For binary outcomes, mean-level change was quantified as the difference of dependent proportions and differential stability as Cohen's kappa.

Baseline factors associated with the course were tested in separate regression models per outcome–predictor pair, each adjusting for the corresponding baseline outcome index (linear regression for continuous outcomes, logistic regression for diagnostic status and functioning outcomes). For IPDE diagnostic outcomes, the corresponding dimensional baseline IPDE score served as the baseline adjustment variable. We report standardized coefficients (outcome and continuous predictors *z*-standardized; binary predictors kept at 0/1) and, for binary outcomes, odds ratios per standard deviation of the predictor. To control for multiple testing, *p* values were adjusted within each analysis family (mean-level tests, stability tests, and the association tests within each outcome) using the Benjamini–Hochberg false discovery rate (FDR; 21). For outcomes with at least two FDR-significant associations, a joint multivariable model including the autoregressor and all FDR-significant predictors was estimated. Because these models were specified on the basis of the FDR screening, their p values should be interpreted descriptively.

Two sets of sensitivity analyses were conducted. First, mean-level change and stability of the continuous outcomes were re-estimated by full-information maximum likelihood with attrition-related auxiliary variables (saturated-correlates approach; 22) using lavaan and semTools (23) (Table S4). Second, we tested the prediction that the follow-up interval relates negatively and non-linearly to the 5-year but not to the 12-month IPDE outcomes, with the BSL-23 and ZAN-BPD, which have no time window mismatch, serving as negative controls. Linear, coverage-based, and restricted-cubic-spline models were compared using the observed data (Table S5).

**Table S1.** *Specification of the Imputation Models*

| Imputation target | *k* | Predictors in the imputation model |
| --- | --- | --- |
| IPDE block (4 summary scores, mpmm) | 12 | BPD criteria (T0), BPD dimensional score (T0), BPD diagnosis (T0), Follow-up interval, Age (T0), Educational attainment (T0), Married/marriage-like relationship (T0), Employed or in education (T0), BSL-23 (T0), ZAN-BPD (T0), DERS (T0), FDS (T0) |
| Age (T0) | 5 | Married/marriage-like relationship (T0), Independent living (T0), Number of biological children (T0), Employed or in education (T1), Follow-up interval |
| Migration background (T0) | 13 | Independent living (T0), DERS (T0), Current eating disorder (T0), Lifetime eating disorder (T0), BSL-23 (T1), BPD diagnosis, 12m (T1), BPD diagnosis, 5y (T1), Stable partnership (T1), BPD criteria, 12m (T1), BPD dimensional score, 12m (T1), BPD criteria, 5y (T1), BPD dimensional score, 5y (T1), Follow-up interval |
| Educational attainment (T0) | 11 | Age (T0), Female sex (T0), Employed or in education (T0), Number of biological children (T0), Outpatient psychotherapy (T0), DERS (T0), Lifetime anxiety disorder (T0), BPD diagnosis, 12m (T1), Outpatient psychotherapy (T1), BPD criteria, 12m (T1), Follow-up interval |
| Married/marriage-like relationship (T0) | 17 | Age (T0), Independent living (T0), Number of biological children (T0), DERS (T0), Current eating disorder (T0), Lifetime eating disorder (T0), BSL-23 (T1), BPD diagnosis, 12m (T1), BPD diagnosis, 5y (T1), Married/marriage-like relationship (T1), Outpatient psychotherapy (T1), Stable partnership (T1), BPD criteria, 12m (T1), BPD dimensional score, 12m (T1), BPD criteria, 5y (T1), BPD dimensional score, 5y (T1), Follow-up interval |
| Employed or in education (T0) | 10 | Female sex (T0), Educational attainment (T0), Outpatient psychotherapy (T0), CTQ-SF (T0), ZAN-BPD (T1), Employed or in education (T1), BPD criteria, 12m (T1), BPD dimensional score, 12m (T1), BPD dimensional score, 5y (T1), Follow-up interval |
| Independent living (T0) | 16 | Age (T0), Married/marriage-like relationship (T0), Number of biological children (T0), DERS (T0), BIS-10 (T0), Current eating disorder (T0), Lifetime eating disorder (T0), BSL-23 (T1), ZAN-BPD (T1), BPD diagnosis, 12m (T1), BPD diagnosis, 5y (T1), BPD criteria, 12m (T1), BPD dimensional score, 12m (T1), BPD criteria, 5y (T1), BPD dimensional score, 5y (T1), Follow-up interval |
| Number of biological children (T0) | 16 | Age (T0), Educational attainment (T0), Married/marriage-like relationship (T0), Independent living (T0), BPD diagnosis (T0), BPD criteria (T0), BPD dimensional score (T0), ZAN-BPD (T0), DERS (T0), BIS-10 (T0), FDS (T0), Lifetime affective disorder (T0), Married/marriage-like relationship (T1), Outpatient psychotherapy (T1), Stable partnership (T1), Follow-up interval |
| Psychopharmacological treatment (T0) | 18 | Female sex (T0), Educational attainment (T0), Number of biological children (T0), Outpatient psychotherapy (T0), DERS (T0), Current eating disorder (T0), Lifetime eating disorder (T0), BSL-23 (T1), BPD diagnosis, 12m (T1), BPD diagnosis, 5y (T1), Psychopharmacological treatment (T1), Outpatient psychotherapy (T1), Stable partnership (T1), BPD criteria, 12m (T1), BPD dimensional score, 12m (T1), BPD criteria, 5y (T1), BPD dimensional score, 5y (T1), Follow-up interval |
| Inpatient psychiatric treatment (T0) | 18 | BPD diagnosis (T0), BPD criteria (T0), BPD dimensional score (T0), ZAN-BPD (T0), DERS (T0), BIS-10 (T0), Current eating disorder (T0), Lifetime eating disorder (T0), BSL-23 (T1), BPD diagnosis, 12m (T1), BPD diagnosis, 5y (T1), Outpatient psychotherapy (T1), Stable partnership (T1), BPD criteria, 12m (T1), BPD dimensional score, 12m (T1), BPD criteria, 5y (T1), BPD dimensional score, 5y (T1), Follow-up interval |
| Outpatient psychotherapy (T0) | 18 | Employed or in education (T0), Psychopharmacological treatment (T0), BPD diagnosis (T0), BPD criteria (T0), BPD dimensional score (T0), DERS (T0), Current eating disorder (T0), Lifetime eating disorder (T0), BSL-23 (T1), BPD diagnosis, 12m (T1), BPD diagnosis, 5y (T1), Outpatient psychotherapy (T1), Stable partnership (T1), BPD criteria, 12m (T1), BPD dimensional score, 12m (T1), BPD criteria, 5y (T1), BPD dimensional score, 5y (T1), Follow-up interval |
| Lifetime suicide attempt (T0) | 16 | Employed or in education (T0), Number of biological children (T0), DERS (T0), CTQ-SF (T0), Current eating disorder (T0), Lifetime eating disorder (T0), BSL-23 (T1), BPD diagnosis, 12m (T1), BPD diagnosis, 5y (T1), Outpatient psychotherapy (T1), Stable partnership (T1), BPD criteria, 12m (T1), BPD dimensional score, 12m (T1), BPD criteria, 5y (T1), BPD dimensional score, 5y (T1), Follow-up interval |
| BSL-23 (T0) | 27 | Educational attainment (T0), Married/marriage-like relationship (T0), Number of biological children (T0), Psychopharmacological treatment (T0), BPD diagnosis (T0), BPD criteria (T0), BPD dimensional score (T0), ZAN-BPD (T0), DERS (T0), BIS-10 (T0), FDS (T0), Current affective disorder (T0), Current anxiety disorder (T0), Current eating disorder (T0), Lifetime eating disorder (T0), BSL-23 (T1), ZAN-BPD (T1), BPD diagnosis, 12m (T1), BPD diagnosis, 5y (T1), Employed or in education (T1), Psychopharmacological treatment (T1), Stable partnership (T1), BPD criteria, 12m (T1), BPD dimensional score, 12m (T1), BPD criteria, 5y (T1), BPD dimensional score, 5y (T1), Follow-up interval |
| ZAN-BPD (T0) | 22 | Inpatient psychiatric treatment (T0), BPD diagnosis (T0), BPD criteria (T0), BPD dimensional score (T0), BSL-23 (T0), DERS (T0), BIS-10 (T0), FDS (T0), Current affective disorder (T0), Current eating disorder (T0), BSL-23 (T1), ZAN-BPD (T1), BPD diagnosis, 12m (T1), BPD diagnosis, 5y (T1), Married/marriage-like relationship (T1), Employed or in education (T1), Outpatient psychotherapy (T1), BPD criteria, 12m (T1), BPD dimensional score, 12m (T1), BPD criteria, 5y (T1), BPD dimensional score, 5y (T1), Follow-up interval |
| DERS (T0) | 25 | Female sex (T0), Independent living (T0), Number of biological children (T0), Outpatient psychotherapy (T0), BPD diagnosis (T0), BPD criteria (T0), BPD dimensional score (T0), BSL-23 (T0), ZAN-BPD (T0), BIS-10 (T0), FDS (T0), Current anxiety disorder (T0), Lifetime eating disorder (T0), BSL-23 (T1), ZAN-BPD (T1), BPD diagnosis, 12m (T1), BPD diagnosis, 5y (T1), Married/marriage-like relationship (T1), Psychopharmacological treatment (T1), Stable partnership (T1), BPD criteria, 12m (T1), BPD dimensional score, 12m (T1), BPD criteria, 5y (T1), BPD dimensional score, 5y (T1), Follow-up interval |
| BIS-10 (T0) | 18 | Independent living (T0), Number of biological children (T0), BPD diagnosis (T0), BPD criteria (T0), BPD dimensional score (T0), BSL-23 (T0), ZAN-BPD (T0), DERS (T0), FDS (T0), Lifetime anxiety disorder (T0), Lifetime eating disorder (T0), BSL-23 (T1), BPD diagnosis, 12m (T1), Psychopharmacological treatment (T1), Stable partnership (T1), BPD criteria, 12m (T1), BPD criteria, 5y (T1), Follow-up interval |
| FDS (T0) | 16 | Independent living (T0), BPD diagnosis (T0), BPD criteria (T0), BPD dimensional score (T0), BSL-23 (T0), ZAN-BPD (T0), DERS (T0), BIS-10 (T0), CTQ-SF (T0), Current anxiety disorder (T0), Lifetime eating disorder (T0), BSL-23 (T1), Stable partnership (T1), BPD criteria, 12m (T1), BPD dimensional score, 5y (T1), Follow-up interval |
| CTQ-SF (T0) | 11 | Educational attainment (T0), Married/marriage-like relationship (T0), Employed or in education (T0), Number of biological children (T0), Lifetime suicide attempt (T0), FDS (T0), Current anxiety disorder (T0), BSL-23 (T1), Psychopharmacological treatment (T1), Stable partnership (T1), Follow-up interval |
| BSL-23 (T1) | 16 | Age (T0), Married/marriage-like relationship (T0), Employed or in education (T0), Psychopharmacological treatment (T0), BPD diagnosis (T0), BPD criteria (T0), BPD dimensional score (T0), BSL-23 (T0), ZAN-BPD (T0), DERS (T0), BIS-10 (T0), FDS (T0), CTQ-SF (T0), Current affective disorder (T0), Current anxiety disorder (T0), Follow-up interval |
| ZAN-BPD (T1) | 11 | Age (T0), Educational attainment (T0), Married/marriage-like relationship (T0), Employed or in education (T0), BPD diagnosis (T0), BPD criteria (T0), BPD dimensional score (T0), BSL-23 (T0), ZAN-BPD (T0), DERS (T0), Follow-up interval |
| BPD diagnosis, 12m (T1) | 11 | Age (T0), Educational attainment (T0), Married/marriage-like relationship (T0), Employed or in education (T0), BPD diagnosis (T0), BPD criteria (T0), BPD dimensional score (T0), BSL-23 (T0), ZAN-BPD (T0), DERS (T0), Follow-up interval |
| BPD diagnosis, 5y (T1) | 11 | Age (T0), Educational attainment (T0), Married/marriage-like relationship (T0), Employed or in education (T0), BPD diagnosis (T0), BPD criteria (T0), BPD dimensional score (T0), BSL-23 (T0), ZAN-BPD (T0), DERS (T0), Follow-up interval |
| Married/marriage-like relationship (T1) | 6 | Age (T0), Married/marriage-like relationship (T0), Employed or in education (T0), BPD diagnosis (T0), DERS (T0), Follow-up interval |
| Employed or in education (T1) | 8 | Age (T0), Married/marriage-like relationship (T0), Employed or in education (T0), BPD diagnosis (T0), BPD criteria (T0), BPD dimensional score (T0), BSL-23 (T0), Follow-up interval |
| Psychopharmacological treatment (T1) | 6 | Age (T0), Married/marriage-like relationship (T0), Employed or in education (T0), Psychopharmacological treatment (T0), BPD diagnosis (T0), Follow-up interval |
| Outpatient psychotherapy (T1) | 7 | Age (T0), Married/marriage-like relationship (T0), Employed or in education (T0), BPD diagnosis (T0), ZAN-BPD (T0), Outpatient psychotherapy (T0), Follow-up interval |
| Stable partnership (T1) | 7 | Age (T0), Married/marriage-like relationship (T0), Employed or in education (T0), Number of biological children (T0), BPD diagnosis (T0), BSL-23 (T0), Follow-up interval |

*Note.* The four IPDE follow-up summary scores were imputed jointly as one block by multivariate predictive mean matching; all other targets were imputed univariately by predictive mean matching. *k* = number of predictors. Predictors were selected per target by association thresholds with the target's observed values (|*r*| ≥ .20) or its missingness indicator (|*r*| ≥ .10), supplemented by the corresponding baseline score and the follow-up interval. BIS = Barratt Impulsiveness Scale; BPD = borderline personality disorder; BSL-23 = Borderline Symptom List, short version; CTQ = Childhood Trauma Questionnaire; DERS = Difficulties in Emotion Regulation Scale; FDS = Fragebogen zu Dissoziativen Symptomen (German version of the Dissociative Experiences Scale, DES); IPDE = International Personality Disorder Examination; MPMM = multivariate predictive mean matching; T0 = baseline; T1 = follow-up; ZAN-BPD = Zanarini Rating Scale for Borderline Personality Disorder; 12m = 12-month assessment window; 5y = 5-year assessment window.T0 = baseline; T1 = follow-up; 12m = 12-month assessment window; 5y = 5-year assessment window. BPD diagnosis, BPD criteria, and BPD dimensional score refer to the IPDE; stable partnership was assessed at follow-up only.

**Supplementary Results**

**Table S2.** *Attrition Analysis: Point-Biserial Correlations of Baseline Variables With Non-Participation at Follow-Up*

| Baseline variable | *r* | *p* | *p*_FDR_ |
| --- | --- | --- | --- |
| Age | -.10 | .043 | .387 |
| Female sex | -.10 | .049 | .387 |
| Migration background | .07 | .163 | .584 |
| Educational attainment | -.09 | .099 | .516 |
| Married or marriage-like relationship | -.11 | .032 | .387 |
| Employed or in education | -.15 | .046 | .387 |
| Independent living | -.07 | .211 | .584 |
| Number of biological children | .03 | .749 | .902 |
| Psychopharmacological treatment | -.08 | .132 | .528 |
| Inpatient psychiatric treatment | .05 | .344 | .704 |
| Outpatient psychotherapy | -.03 | .547 | .779 |
| Lifetime suicide attempt | -.04 | .452 | .751 |
| BPD diagnosis (IPDE) | .10 | .041 | .387 |
| Number of BPD criteria (IPDE) | .06 | .229 | .598 |
| Dimensional BPD score (IPDE) | .07 | .180 | .584 |
| BPD symptom severity (BSL-23) | -.00 | .948 | .950 |
| BPD symptom severity (ZAN-BPD) | .05 | .296 | .704 |
| Emotion dysregulation (DERS) | -.01 | .911 | .950 |
| Impulsivity (BIS-10) | -.01 | .859 | .939 |
| Dissociation (DES) | .03 | .569 | .786 |
| Childhood maltreatment (CTQ-SF) | .08 | .116 | .528 |
| Current affective disorder | -.00 | .950 | .950 |
| Lifetime affective disorder | -.06 | .210 | .584 |
| Lifetime psychotic disorder | -.02 | .744 | .902 |
| Lifetime substance use disorder | -.02 | .735 | .902 |
| Current anxiety disorder | .03 | .603 | .809 |
| Lifetime anxiety disorder | -.05 | .335 | .704 |
| Current somatoform disorder | -.01 | .858 | .939 |
| Current eating disorder | .01 | .824 | .939 |
| Lifetime eating disorder | -.06 | .200 | .584 |
| Lifetime adjustment or other disorder | -.03 | .536 | .779 |
| Follow-up interval | .16 | .001 | .053 |

*Note.* Positive correlations indicate that higher values were associated with non-participation at follow-up. *p*_FDR_ = Benjamini–Hochberg-adjusted *p* value. No association remained significant after FDR correction across all 47 baseline variables tested (including 15 additional baseline instruments not listed here); the follow-up interval showed the numerically largest association (*r* = .16). BIS-10 = Barratt Impulsiveness Scale, Version 10; BPD = borderline personality disorder; BSL-23 = Borderline Symptom List, short version; CTQ = Childhood Trauma Questionnaire Short Form; DERS = Difficulties in Emotion Regulation Scale; FDS = Fragebogen zu Dissoziativen Symptomen (German version of the Dissociative Experiences Scale [DES]); IPDE = International Personality Disorder Examination; ZAN-BPD = Zanarini Rating Scale for Borderline Personality Disorder.

**Table S3.** *Multiple Imputation Quality Check: Observed, Imputed, and Pooled Means, and Observed Versus Pooled Stability Estimates*

| Variable | *M* observed | *M* imputed | *M* pooled | \|diff\| / *SD* |
| --- | --- | --- | --- | --- |
| Age (baseline) | 28.799 | 28.295 | 28.797 | 0.06 |
| Female sex (baseline) | 0.861 | — | 0.861 | — |
| Migration background (baseline) | 0.031 | 0.059 | 0.036 | 0.16 |
| Educational attainment (baseline) | 2.440 | 2.397 | 2.432 | 0.06 |
| BPD diagnosis (IPDE) (baseline) | 0.841 | — | 0.841 | — |
| Number of BPD criteria (IPDE) (baseline) | 5.527 | — | 5.527 | — |
| Dimensional BPD score (IPDE) (baseline) | 12.829 | — | 12.829 | — |
| BPD symptom severity (BSL-23) (baseline) | 1.658 | 1.596 | 1.650 | 0.07 |
| BPD symptom severity (ZAN-BPD) (baseline) | 1.212 | 1.398 | 1.216 | 0.28 |
| Married or marriage-like relationship (baseline) | 0.223 | 0.222 | 0.223 | 0 |
| Employed or in education (baseline) | 0.783 | 0.757 | 0.768 | 0.06 |
| Independent living (baseline) | 0.827 | 0.798 | 0.822 | 0.08 |
| Number of biological children (baseline) | 0.671 | 0.576 | 0.611 | 0.09 |
| Psychopharmacological treatment (baseline) | 0.214 | 0.202 | 0.213 | 0.03 |
| Inpatient psychiatric treatment (baseline) | 0.314 | 0.327 | 0.316 | 0.03 |
| Outpatient psychotherapy (baseline) | 0.595 | 0.587 | 0.594 | 0.02 |
| Lifetime suicide attempt (baseline) | 0.556 | 0.572 | 0.558 | 0.03 |
| Emotion dysregulation (DERS) (baseline) | 120.289 | 121.652 | 120.455 | 0.05 |
| Impulsivity (BIS-10) (baseline) | 81.809 | 83.105 | 81.960 | 0.1 |
| Dissociation (DES) (baseline) | 18.633 | 19.225 | 18.702 | 0.05 |
| Childhood maltreatment (CTQ) (baseline) | 61.109 | 61.694 | 61.200 | 0.03 |
| Current affective disorder (baseline) | 0.305 | — | 0.305 | — |
| Lifetime affective disorder (baseline) | 0.795 | — | 0.795 | — |
| Lifetime psychotic disorder (baseline) | 0.029 | — | 0.029 | — |
| Lifetime substance use disorder (baseline) | 0.234 | — | 0.234 | — |
| Current anxiety disorder (baseline) | 0.520 | — | 0.520 | — |
| Lifetime anxiety disorder (baseline) | 0.663 | — | 0.663 | — |
| Current somatoform disorder (baseline) | 0.046 | — | 0.046 | — |
| Current eating disorder (baseline) | 0.215 | — | 0.215 | — |
| Lifetime eating disorder (baseline) | 0.444 | — | 0.444 | — |
| Lifetime adjustment or other disorder (baseline) | 0.027 | — | 0.027 | — |
| BPD diagnosis (IPDE), 12-month window (follow-up) | 0.405 | 0.364 | 0.382 | 0.08 |
| BPD diagnosis (IPDE), 5-year window (follow-up) | 0.551 | 0.618 | 0.588 | 0.13 |
| Number of BPD criteria (IPDE), 12-month window (follow-up) | 3.832 | 3.867 | 3.851 | 0.01 |
| Number of BPD criteria (IPDE), 5-year window (follow-up) | 4.654 | 4.895 | 4.787 | 0.09 |
| Dimensional BPD score (IPDE), 12-month window (follow-up) | 9.870 | 10.072 | 9.981 | 0.04 |
| Dimensional BPD score (IPDE), 5-year window (follow-up) | 11.314 | 11.790 | 11.575 | 0.1 |
| BPD symptom severity (BSL-23) (follow-up) | 1.320 | 1.294 | 1.305 | 0.03 |
| BPD symptom severity (ZAN-BPD) (follow-up) | 0.861 | 0.851 | 0.856 | 0.01 |
| Married or marriage-like relationship (follow-up) | 0.328 | 0.304 | 0.315 | 0.05 |
| In stable relationship (follow-up)^a^ | 0.585 | 0.578 | 0.581 | 0.01 |
| Employed or in education (follow-up) | 0.683 | 0.685 | 0.684 | 0 |
| Psychopharmacological treatment (follow-up) | 0.437 | 0.420 | 0.428 | 0.03 |
| Outpatient psychotherapy (follow-up) | 0.415 | 0.403 | 0.409 | 0.02 |

| Outcome | Stability (observed) | Stability (pooled) |
| --- | --- | --- |
| BPD diagnosis (IPDE), 12-month window | .28 | .23 |
| BPD diagnosis (IPDE), 5-year window | .31 | .28 |
| Number of BPD criteria (IPDE), 12-month window | .40 | .38 |
| Number of BPD criteria (IPDE), 5-year window | .45 | .41 |
| Dimensional BPD score (IPDE), 12-month window | .41 | .40 |
| Dimensional BPD score (IPDE), 5-year window | .47 | .43 |
| BPD symptom severity (BSL-23) | .59 | .54 |
| BPD symptom severity (ZAN-BPD) | .37 | .30 |
| Married or marriage-like relationship | .40 | .37 |
| Employed or in education | .22 | .33 |
| Psychopharmacological treatment | .31 | .26 |
| Outpatient psychotherapy | .18 | .17 |

*Note.* Upper panel: means of observed values, imputed values (average across the 100 imputations, imputed cases only), and pooled complete-data estimates; |diff| / *SD* = absolute standardized difference between observed and imputed means. — = variable fully observed (nothing imputed). Differences ≤ 0.30 *SD* were considered unremarkable given predicted mean-level differences between completers and non-completers. Lower panel: differential stability (Pearson correlation; for binary outcomes the phi coefficient, which differs from the κ reported in Table 2) estimated on the observed data versus pooled across imputations. BIS-10, Barratt Impulsiveness Scale, Version 10; BPD, borderline personality disorder; BSL-23, Borderline Symptom List, short version; CTQ, Childhood Trauma Questionnaire; DERS, Difficulties in Emotion Regulation Scale; DES, Dissociative Experiences Scale; IPDE, International Personality Disorder Examination; *M*, mean; *SD*, standard deviation; ZAN-BPD, Zanarini Rating Scale for Borderline Personality Disorder.

^a^ only assessed at follow-up

*Sensitivity Analyses of Longitudinal Change and Stability*

Two sets of sensitivity analyses were conducted to evaluate the robustness of the primary findings and the potential influence of assessment-window coverage. First, full-information maximum likelihood estimates of change and stability were similar to the multiple-imputation estimates (e.g., number of criteria, 12-month window: difference = −1.65 vs. −1.68; *r* = .37 vs. .38; Table S4).

**Table S4.** *Sensitivity Analysis: Full-Information Maximum Likelihood (Saturated-Correlates Approach) Versus Multiple Imputation for Continuous Outcomes*

| Outcome | FIML T0 | FIML T1 | FIML Δ | *p* | FIML *r* | MI Δ | MI *r* |
| --- | --- | --- | --- | --- | --- | --- | --- |
| Number of BPD criteria (IPDE), 12-month window | 5.53 | 3.88 | -1.65 | <.001 | .37 | -1.68 | .38 |
| Number of BPD criteria (IPDE), 5-year window | 5.53 | 4.64 | -0.89 | <.001 | .42 | -0.74 | .41 |
| Dimensional BPD score (IPDE), 12-month window | 12.83 | 10.00 | -2.83 | <.001 | .39 | -2.85 | .40 |
| Dimensional BPD score (IPDE), 5-year window | 12.83 | 11.38 | -1.45 | <.001 | .43 | -1.25 | .43 |
| BPD symptom severity  (BSL-23) | 1.66 | 1.31 | -0.34 | <.001 | .57 | -0.34 | .54 |
| BPD symptom severity  (ZAN-BPD) | 1.21 | 0.89 | -0.32 | <.001 | .35 | -0.36 | .30 |

*Note.* FIML models included six attrition-related auxiliary variables (saturated-correlates approach). Δ = mean difference between occasions; *r* = differential stability. MI columns repeat the primary multiple-imputation estimates from Table 2 for comparison. BPD, borderline personality disorder; BSL-23, Borderline Symptom List, short version; FIML, full-information maximum likelihood; IPDE, International Personality Disorder Examination; MI, multiple imputation; T0, baseline; T1, follow-up; ZAN-BPD, Zanarini Rating Scale for Borderline Personality Disorder.

Second, consistent with the predicted window-coverage mechanism in IPDE scores, a longer follow-up interval was associated with lower symptom levels in the 5-year window (number of criteria: *r* = −.24, *p* < .001; non-linear spline test, *p* = .003) but not in the 12-month window (*r* = −.11, *p* = .138). No linear associations were observed for the BSL-23 or ZAN-BPD (|*r*| ≤ .09, *p* ≥ .204), although the spline test indicated a non-linear association for the BSL-23 (*p* = .037) (Table S5; unadjusted *p* values).

**Table S5.** *Sensitivity Analysis: Association of the Follow-Up Interval With Outcome Levels at Follow-Up (Observed Data)*

| Outcome | *r*  interval | *p* interval | *b*  linear | *p*  linear | *b*  coverage | *p*  coverage | *p*  spline | *b* moderation | *p* moderation |
| --- | --- | --- | --- | --- | --- | --- | --- | --- | --- |
| Number of BPD criteria  (IPDE), 12-month window | -.11 | .138 | -0.008 | .930 | 0.186 | .877 | .600 | -0.046 | .266 |
| Number of BPD criteria  (IPDE), 5-year window | -.24 | <.001 | -0.189 | .051 | 2.682 | .028 | .003 | -0.069 | .101 |
| Dimensional BPD score  (IPDE), 12-month window | -.14 | .053 | -0.065 | .720 | 1.168 | .609 | .235 | -0.050 | .246 |
| Dimensional BPD score  (IPDE), 5-year window | -.23 | .002 | -0.271 | .125 | 4.027 | .072 | .004 | -0.077 | .065 |
| BPD symptom severity  (BSL-23) | -.09 | .204 | -0.046 | .164 | 0.608 | .152 | .037 | -0.036 | .306 |
| BPD symptom severity  (ZAN-BPD) | -.09 | .228 | -0.013 | .647 | 0.206 | .553 | .539 | 0.011 | .793 |

*Note. r* interval = correlation of the follow-up interval with the outcome at follow-up; linear, coverage-based, and restricted-cubic-spline models were compared, and moderation refers to interval × baseline interaction. Consistent with the window-coverage mechanism, the interval related negatively and non-linearly to the 5-year IPDE outcomes but not to the 12-month IPDE outcomes; for the BSL-23 and ZAN-BPD, which have no time-window mismatch, no linear association emerged, although the spline test was nominally significant for the BSL-23. BPD, borderline personality disorder; BSL-23, Borderline Symptom List, short version; IPDE, International Personality Disorder Examination; ZAN-BPD, Zanarini Rating Scale for Borderline Personality Disorder.

**Table S6.** *Complete Bivariate Prediction Models: Baseline Predictors of Outcomes at Follow-Up (Each Model Adjusted for the Baseline Level of the Outcome)*

| Outcome | Baseline predictor | *b* (SE) | 95% CI | β / OR_SD_ [95% CI] | *p* | *p*_FDR_ |
| --- | --- | --- | --- | --- | --- | --- |
| BPD diagnosis (IPDE),  12-month window | Age | 0.034 (0.018) | [-0.001, 0.069] | 1.32 [0.99, 1.76] | .054 | .215 |
|  | Female sex | 0.170 (0.385) | [-0.590, 0.931] | 1.19 [0.55, 2.54] | .659 | .823 |
|  | Migration background | 0.452 (0.811) | [-1.152, 2.056] | 1.57 [0.32, 7.82] | .578 | .809 |
|  | Educational attainment | -0.368 (0.201) | [-0.765, 0.029] | 0.77 [0.58, 1.02] | .069 | .215 |
|  | Married or marriage-like relationship | 0.876 (0.334) | [0.217, 1.536] | 2.40 [1.24, 4.64] | .009 | .063 |
|  | Employed or in education | -0.593 (0.442) | [-1.473, 0.288] | 0.55 [0.23, 1.33] | .184 | .429 |
|  | Independent living | -0.119 (0.362) | [-0.835, 0.597] | 0.89 [0.43, 1.82] | .743 | .823 |
|  | Number of biological children | 0.278 (0.147) | [-0.012, 0.568] | 1.30 [0.99, 1.70] | .061 | .215 |
|  | Psychopharmacological treatment | 0.091 (0.303) | [-0.506, 0.688] | 1.09 [0.60, 1.99] | .764 | .823 |
|  | Inpatient psychiatric treatment | -0.143 (0.303) | [-0.741, 0.455] | 0.87 [0.48, 1.58] | .637 | .823 |
|  | Outpatient psychotherapy | 0.184 (0.274) | [-0.357, 0.725] | 1.20 [0.70, 2.07] | .502 | .740 |
|  | Lifetime suicide attempt | 0.104 (0.268) | [-0.424, 0.632] | 1.11 [0.65, 1.88] | .699 | .823 |
|  | BPD symptom severity (BSL-23) | 0.940 (0.212) | [0.521, 1.359] | 2.33 [1.60, 3.40] | <.001 | <.001 |
|  | BPD symptom severity (ZAN-BPD) | 0.664 (0.259) | [0.153, 1.175] | 1.55 [1.11, 2.19] | .011 | .063 |
|  | Emotion dysregulation (DERS) | 0.034 (0.008) | [0.019, 0.050] | 2.34 [1.58, 3.45] | <.001 | <.001 |
|  | Impulsivity (BIS-10) | 0.017 (0.010) | [-0.003, 0.037] | 1.26 [0.96, 1.65] | .099 | .277 |
|  | Dissociation (DES) | 0.019 (0.011) | [-0.001, 0.040] | 1.27 [0.98, 1.66] | .069 | .215 |
|  | Childhood maltreatment (CTQ) | 0.018 (0.007) | [0.005, 0.031] | 1.42 [1.09, 1.83] | .008 | .063 |
|  | Current affective disorder | 0.253 (0.275) | [-0.288, 0.795] | 1.29 [0.75, 2.21] | .357 | .667 |
|  | Lifetime affective disorder | 0.123 (0.334) | [-0.536, 0.781] | 1.13 [0.58, 2.18] | .714 | .823 |
|  | Lifetime psychotic disorder | — ^a^ | — | — | .989 | .991 |
|  | Lifetime substance use disorder | 0.250 (0.302) | [-0.346, 0.845] | 1.28 [0.71, 2.33] | .410 | .675 |
|  | Current anxiety disorder | 0.315 (0.262) | [-0.201, 0.831] | 1.37 [0.82, 2.30] | .230 | .460 |
|  | Lifetime anxiety disorder | 0.410 (0.281) | [-0.144, 0.964] | 1.51 [0.87, 2.62] | .146 | .371 |
|  | Current somatoform disorder | -0.503 (0.727) | [-1.939, 0.933] | 0.60 [0.14, 2.54] | .490 | .740 |
|  | Current eating disorder | 0.257 (0.302) | [-0.339, 0.853] | 1.29 [0.71, 2.35] | .396 | .675 |
|  | Lifetime eating disorder | 0.305 (0.251) | [-0.190, 0.800] | 1.36 [0.83, 2.23] | .226 | .460 |
|  | Lifetime adjustment or other disorder | — ^a^ | — | — | .991 | .991 |
| BPD diagnosis (IPDE),  5-year window | Age | 0.025 (0.017) | [-0.010, 0.059] | 1.23 [0.92, 1.62] | .158 | .402 |
|  | Female sex | 0.075 (0.400) | [-0.715, 0.864] | 1.08 [0.49, 2.37] | .852 | .898 |
|  | Migration background | 0.335 (0.880) | [-1.406, 2.076] | 1.40 [0.24, 7.97] | .704 | .898 |
|  | Educational attainment | -0.336 (0.205) | [-0.742, 0.070] | 0.79 [0.59, 1.05] | .104 | .402 |
|  | Married or marriage-like relationship | 0.668 (0.369) | [-0.061, 1.398] | 1.95 [0.94, 4.05] | .072 | .397 |
|  | Employed or in education | -0.566 (0.462) | [-1.484, 0.352] | 0.57 [0.23, 1.42] | .223 | .417 |
|  | Independent living | -0.064 (0.382) | [-0.819, 0.690] | 0.94 [0.44, 2.00] | .866 | .898 |
|  | Number of biological children | 0.244 (0.161) | [-0.073, 0.561] | 1.26 [0.93, 1.69] | .130 | .402 |
|  | Psychopharmacological treatment | 0.238 (0.325) | [-0.403, 0.878] | 1.27 [0.67, 2.41] | .465 | .685 |
|  | Inpatient psychiatric treatment | 0.063 (0.316) | [-0.562, 0.688] | 1.06 [0.57, 1.99] | .842 | .898 |
|  | Outpatient psychotherapy | 0.244 (0.281) | [-0.311, 0.800] | 1.28 [0.73, 2.23] | .386 | .636 |
|  | Lifetime suicide attempt | -0.085 (0.278) | [-0.634, 0.464] | 0.92 [0.53, 1.59] | .761 | .898 |
|  | BPD symptom severity (BSL-23) | 0.891 (0.214) | [0.468, 1.313] | 2.23 [1.52, 3.26] | <.001 | <.001 |
|  | BPD symptom severity (ZAN-BPD) | 0.696 (0.269) | [0.164, 1.228] | 1.59 [1.12, 2.26] | .011 | .075 |
|  | Emotion dysregulation (DERS) | 0.033 (0.008) | [0.018, 0.048] | 2.25 [1.55, 3.25] | <.001 | <.001 |
|  | Impulsivity (BIS-10) | 0.017 (0.011) | [-0.004, 0.038] | 1.25 [0.94, 1.67] | .119 | .402 |
|  | Dissociation (DES) | 0.020 (0.012) | [-0.003, 0.044] | 1.29 [0.96, 1.73] | .085 | .397 |
|  | Childhood maltreatment (CTQ) | 0.019 (0.007) | [0.005, 0.033] | 1.44 [1.11, 1.89] | .007 | .065 |
|  | Current affective disorder | 0.416 (0.297) | [-0.170, 1.003] | 1.52 [0.84, 2.73] | .163 | .402 |
|  | Lifetime affective disorder | 0.087 (0.335) | [-0.575, 0.749] | 1.09 [0.56, 2.11] | .797 | .898 |
|  | Lifetime psychotic disorder | -0.644 (0.818) | [-2.258, 0.970] | 0.53 [0.10, 2.64] | .432 | .672 |
|  | Lifetime substance use disorder | 0.111 (0.308) | [-0.496, 0.718] | 1.12 [0.61, 2.05] | .719 | .898 |
|  | Current anxiety disorder | 0.381 (0.278) | [-0.168, 0.929] | 1.46 [0.85, 2.53] | .172 | .402 |
|  | Lifetime anxiety disorder | 0.369 (0.291) | [-0.205, 0.943] | 1.45 [0.81, 2.57] | .206 | .412 |
|  | Current somatoform disorder | -0.077 (0.654) | [-1.367, 1.213] | 0.93 [0.26, 3.36] | .906 | .906 |
|  | Current eating disorder | 0.391 (0.331) | [-0.262, 1.043] | 1.48 [0.77, 2.84] | .239 | .418 |
|  | Lifetime eating disorder | 0.347 (0.266) | [-0.177, 0.871] | 1.42 [0.84, 2.39] | .193 | .412 |
|  | Lifetime adjustment or other disorder | -0.243 (0.808) | [-1.836, 1.350] | 0.78 [0.16, 3.86] | .764 | .898 |
| Number of BPD criteria (IPDE), 12-month window | Age | 0.033 (0.018) | [-0.002, 0.068] | 0.12 [-0.01, 0.24] | .068 | .375 |
|  | Female sex | -0.089 (0.414) | [-0.907, 0.729] | -0.04 [-0.40, 0.32] | .829 | .957 |
|  | Migration background | 0.122 (0.872) | [-1.606, 1.851] | 0.05 [-0.71, 0.81] | .889 | .957 |
|  | Educational attainment | -0.504 (0.211) | [-0.922, -0.086] | -0.16 [-0.29, -0.03] | .018 | .130 |
|  | Married or marriage-like relationship | 0.474 (0.324) | [-0.166, 1.113] | 0.21 [-0.07, 0.49] | .146 | .453 |
|  | Employed or in education | -0.720 (0.475) | [-1.667, 0.227] | -0.32 [-0.73, 0.10] | .134 | .453 |
|  | Independent living | -0.096 (0.396) | [-0.879, 0.687] | -0.04 [-0.39, 0.30] | .809 | .957 |
|  | Number of biological children | 0.258 (0.146) | [-0.032, 0.547] | 0.11 [-0.01, 0.22] | .080 | .375 |
|  | Psychopharmacological treatment | -0.046 (0.326) | [-0.688, 0.596] | -0.02 [-0.30, 0.26] | .888 | .957 |
|  | Inpatient psychiatric treatment | -0.078 (0.326) | [-0.722, 0.566] | -0.03 [-0.32, 0.25] | .812 | .957 |
|  | Outpatient psychotherapy | 0.082 (0.288) | [-0.486, 0.650] | 0.04 [-0.21, 0.29] | .776 | .957 |
|  | Lifetime suicide attempt | 0.056 (0.286) | [-0.509, 0.621] | 0.02 [-0.22, 0.27] | .845 | .957 |
|  | BPD symptom severity (BSL-23) | 0.867 (0.199) | [0.473, 1.261] | 0.34 [0.19, 0.49] | <.001 | <.001 |
|  | BPD symptom severity (ZAN-BPD) | 0.389 (0.294) | [-0.192, 0.971] | 0.11 [-0.06, 0.28] | .188 | .478 |
|  | Emotion dysregulation (DERS) | 0.026 (0.007) | [0.012, 0.040] | 0.28 [0.13, 0.43] | <.001 | .006 |
|  | Impulsivity (BIS-10) | 0.012 (0.011) | [-0.009, 0.033] | 0.07 [-0.05, 0.20] | .257 | .553 |
|  | Dissociation (DES) | 0.013 (0.011) | [-0.008, 0.034] | 0.07 [-0.04, 0.19] | .216 | .503 |
|  | Childhood maltreatment (CTQ) | 0.017 (0.007) | [0.004, 0.030] | 0.15 [0.04, 0.26] | .009 | .085 |
|  | Current affective disorder | 0.253 (0.298) | [-0.336, 0.842] | 0.11 [-0.15, 0.37] | .398 | .796 |
|  | Lifetime affective disorder | 0.015 (0.338) | [-0.652, 0.682] | 0.01 [-0.29, 0.30] | .965 | .983 |
|  | Lifetime psychotic disorder | -0.016 (0.754) | [-1.503, 1.471] | -0.01 [-0.66, 0.65] | .983 | .983 |
|  | Lifetime substance use disorder | 0.111 (0.312) | [-0.504, 0.726] | 0.05 [-0.22, 0.32] | .722 | .957 |
|  | Current anxiety disorder | 0.147 (0.278) | [-0.401, 0.695] | 0.06 [-0.18, 0.30] | .597 | .957 |
|  | Lifetime anxiety disorder | 0.180 (0.281) | [-0.373, 0.734] | 0.08 [-0.16, 0.32] | .521 | .912 |
|  | Current somatoform disorder | -0.240 (0.624) | [-1.470, 0.990] | -0.10 [-0.65, 0.43] | .701 | .957 |
|  | Current eating disorder | 0.438 (0.313) | [-0.180, 1.056] | 0.19 [-0.08, 0.46] | .164 | .459 |
|  | Lifetime eating disorder | 0.411 (0.254) | [-0.091, 0.912] | 0.18 [-0.04, 0.40] | .108 | .432 |
|  | Lifetime adjustment or other disorder | -0.546 (0.767) | [-2.057, 0.964] | -0.24 [-0.90, 0.42] | .477 | .890 |
| Number of BPD criteria (IPDE), 5-year window | Age | 0.024 (0.018) | [-0.012, 0.059] | 0.08 [-0.04, 0.20] | .194 | .536 |
|  | Female sex | -0.091 (0.430) | [-0.941, 0.760] | -0.04 [-0.39, 0.31] | .834 | .921 |
|  | Migration background | -0.162 (0.886) | [-1.918, 1.593] | -0.07 [-0.79, 0.66] | .855 | .921 |
|  | Educational attainment | -0.364 (0.214) | [-0.787, 0.059] | -0.11 [-0.23, 0.02] | .091 | .536 |
|  | Married or marriage-like relationship | 0.472 (0.348) | [-0.215, 1.159] | 0.19 [-0.09, 0.48] | .177 | .536 |
|  | Employed or in education | -0.621 (0.461) | [-1.538, 0.297] | -0.26 [-0.64, 0.12] | .182 | .536 |
|  | Independent living | -0.043 (0.418) | [-0.872, 0.785] | -0.02 [-0.36, 0.32] | .918 | .921 |
|  | Number of biological children | 0.207 (0.153) | [-0.095, 0.510] | 0.08 [-0.04, 0.20] | .178 | .536 |
|  | Psychopharmacological treatment | 0.042 (0.351) | [-0.652, 0.735] | 0.02 [-0.27, 0.30] | .906 | .921 |
|  | Inpatient psychiatric treatment | 0.054 (0.356) | [-0.652, 0.759] | 0.02 [-0.27, 0.31] | .880 | .921 |
|  | Outpatient psychotherapy | 0.187 (0.315) | [-0.436, 0.811] | 0.08 [-0.18, 0.34] | .553 | .886 |
|  | Lifetime suicide attempt | -0.187 (0.299) | [-0.779, 0.405] | -0.08 [-0.32, 0.17] | .534 | .886 |
|  | BPD symptom severity (BSL-23) | 0.747 (0.202) | [0.348, 1.146] | 0.28 [0.13, 0.42] | <.001 | .007 |
|  | BPD symptom severity (ZAN-BPD) | 0.356 (0.293) | [-0.223, 0.934] | 0.10 [-0.06, 0.26] | .226 | .536 |
|  | Emotion dysregulation (DERS) | 0.026 (0.007) | [0.012, 0.040] | 0.27 [0.12, 0.41] | <.001 | .007 |
|  | Impulsivity (BIS-10) | 0.011 (0.012) | [-0.013, 0.034] | 0.06 [-0.07, 0.19] | .365 | .785 |
|  | Dissociation (DES) | 0.014 (0.012) | [-0.009, 0.037] | 0.07 [-0.05, 0.19] | .230 | .536 |
|  | Childhood maltreatment (CTQ) | 0.017 (0.007) | [0.002, 0.031] | 0.13 [0.02, 0.25] | .024 | .222 |
|  | Current affective disorder | 0.213 (0.311) | [-0.402, 0.827] | 0.09 [-0.17, 0.34] | .495 | .886 |
|  | Lifetime affective disorder | -0.046 (0.367) | [-0.772, 0.679] | -0.02 [-0.32, 0.28] | .899 | .921 |
|  | Lifetime psychotic disorder | 0.196 (0.805) | [-1.391, 1.783] | 0.08 [-0.57, 0.74] | .808 | .921 |
|  | Lifetime substance use disorder | -0.033 (0.328) | [-0.680, 0.615] | -0.01 [-0.28, 0.25] | .921 | .921 |
|  | Current anxiety disorder | 0.154 (0.294) | [-0.426, 0.734] | 0.06 [-0.18, 0.30] | .601 | .886 |
|  | Lifetime anxiety disorder | 0.191 (0.302) | [-0.406, 0.788] | 0.08 [-0.17, 0.33] | .528 | .886 |
|  | Current somatoform disorder | -0.119 (0.672) | [-1.445, 1.207] | -0.05 [-0.60, 0.50] | .860 | .921 |
|  | Current eating disorder | 0.396 (0.323) | [-0.240, 1.032] | 0.16 [-0.10, 0.43] | .221 | .536 |
|  | Lifetime eating disorder | 0.405 (0.277) | [-0.141, 0.952] | 0.17 [-0.06, 0.39] | .145 | .536 |
|  | Lifetime adjustment or other disorder | -0.445 (0.820) | [-2.061, 1.171] | -0.18 [-0.85, 0.48] | .588 | .886 |
| Dimensional BPD score (IPDE), 12-month window | Age | 0.047 (0.033) | [-0.018, 0.113] | 0.09 [-0.03, 0.21] | .155 | .477 |
|  | Female sex | -0.368 (0.785) | [-1.919, 1.184] | -0.09 [-0.44, 0.27] | .640 | .916 |
|  | Migration background | 0.423 (1.591) | [-2.728, 3.575] | 0.10 [-0.63, 0.83] | .791 | .916 |
|  | Educational attainment | -0.932 (0.384) | [-1.691, -0.172] | -0.15 [-0.28, -0.03] | .017 | .118 |
|  | Married or marriage-like relationship | 0.845 (0.614) | [-0.367, 2.057] | 0.20 [-0.08, 0.47] | .170 | .477 |
|  | Employed or in education | -1.281 (0.864) | [-3.003, 0.442] | -0.30 [-0.69, 0.10] | .143 | .477 |
|  | Independent living | -0.553 (0.762) | [-2.062, 0.955] | -0.13 [-0.48, 0.22] | .469 | .821 |
|  | Number of biological children | 0.429 (0.272) | [-0.107, 0.966] | 0.09 [-0.02, 0.21] | .116 | .477 |
|  | Psychopharmacological treatment | 0.101 (0.627) | [-1.136, 1.339] | 0.02 [-0.26, 0.31] | .872 | .916 |
|  | Inpatient psychiatric treatment | -0.046 (0.611) | [-1.253, 1.162] | -0.01 [-0.29, 0.27] | .940 | .940 |
|  | Outpatient psychotherapy | 0.169 (0.563) | [-0.943, 1.281] | 0.04 [-0.22, 0.30] | .765 | .916 |
|  | Lifetime suicide attempt | 0.090 (0.538) | [-0.973, 1.153] | 0.02 [-0.22, 0.27] | .868 | .916 |
|  | BPD symptom severity (BSL-23) | 1.617 (0.379) | [0.865, 2.370] | 0.34 [0.19, 0.49] | <.001 | .001 |
|  | BPD symptom severity (ZAN-BPD) | 0.782 (0.539) | [-0.283, 1.848] | 0.12 [-0.04, 0.29] | .149 | .477 |
|  | Emotion dysregulation (DERS) | 0.045 (0.014) | [0.018, 0.072] | 0.26 [0.10, 0.41] | .001 | .019 |
|  | Impulsivity (BIS-10) | 0.018 (0.021) | [-0.023, 0.059] | 0.06 [-0.07, 0.19] | .384 | .767 |
|  | Dissociation (DES) | 0.024 (0.020) | [-0.015, 0.063] | 0.07 [-0.04, 0.18] | .231 | .538 |
|  | Childhood maltreatment (CTQ) | 0.030 (0.013) | [0.005, 0.055] | 0.14 [0.03, 0.24] | .017 | .118 |
|  | Current affective disorder | 0.532 (0.544) | [-0.542, 1.606] | 0.12 [-0.13, 0.37] | .330 | .710 |
|  | Lifetime affective disorder | -0.262 (0.630) | [-1.506, 0.982] | -0.06 [-0.35, 0.23] | .678 | .916 |
|  | Lifetime psychotic disorder | -0.288 (1.415) | [-3.077, 2.501] | -0.07 [-0.71, 0.58] | .839 | .916 |
|  | Lifetime substance use disorder | 0.087 (0.592) | [-1.082, 1.256] | 0.02 [-0.25, 0.29] | .883 | .916 |
|  | Current anxiety disorder | 0.219 (0.525) | [-0.818, 1.256] | 0.05 [-0.19, 0.29] | .677 | .916 |
|  | Lifetime anxiety disorder | 0.168 (0.542) | [-0.902, 1.238] | 0.04 [-0.21, 0.29] | .757 | .916 |
|  | Current somatoform disorder | -0.215 (1.177) | [-2.537, 2.107] | -0.05 [-0.59, 0.49] | .855 | .916 |
|  | Current eating disorder | 0.719 (0.590) | [-0.443, 1.882] | 0.17 [-0.10, 0.44] | .224 | .538 |
|  | Lifetime eating disorder | 0.691 (0.485) | [-0.267, 1.648] | 0.16 [-0.06, 0.38] | .156 | .477 |
|  | Lifetime adjustment or other disorder | -1.147 (1.444) | [-3.992, 1.698] | -0.27 [-0.93, 0.39] | .428 | .798 |
| Dimensional BPD score (IPDE), 5-year window | Age | 0.035 (0.033) | [-0.030, 0.099] | 0.06 [-0.06, 0.18] | .290 | .712 |
|  | Female sex | -0.209 (0.773) | [-1.736, 1.319] | -0.05 [-0.40, 0.30] | .788 | .959 |
|  | Migration background | -0.019 (1.577) | [-3.141, 3.103] | -0.00 [-0.71, 0.70] | .991 | .991 |
|  | Educational attainment | -0.645 (0.377) | [-1.390, 0.100] | -0.10 [-0.22, 0.02] | .089 | .624 |
|  | Married or marriage-like relationship | 0.794 (0.626) | [-0.442, 2.030] | 0.18 [-0.10, 0.46] | .206 | .712 |
|  | Employed or in education | -1.086 (0.808) | [-2.693, 0.520] | -0.25 [-0.61, 0.12] | .182 | .712 |
|  | Independent living | -0.409 (0.777) | [-1.948, 1.131] | -0.09 [-0.44, 0.26] | .600 | .933 |
|  | Number of biological children | 0.317 (0.279) | [-0.234, 0.868] | 0.07 [-0.05, 0.19] | .258 | .712 |
|  | Psychopharmacological treatment | 0.195 (0.651) | [-1.091, 1.480] | 0.04 [-0.25, 0.34] | .765 | .959 |
|  | Inpatient psychiatric treatment | 0.081 (0.628) | [-1.162, 1.324] | 0.02 [-0.26, 0.30] | .898 | .991 |
|  | Outpatient psychotherapy | 0.321 (0.583) | [-0.832, 1.473] | 0.07 [-0.19, 0.34] | .583 | .933 |
|  | Lifetime suicide attempt | -0.314 (0.544) | [-1.389, 0.760] | -0.07 [-0.32, 0.17] | .564 | .933 |
|  | BPD symptom severity (BSL-23) | 1.349 (0.369) | [0.619, 2.080] | 0.28 [0.13, 0.42] | <.001 | .011 |
|  | BPD symptom severity (ZAN-BPD) | 0.619 (0.516) | [-0.400, 1.638] | 0.09 [-0.06, 0.25] | .232 | .712 |
|  | Emotion dysregulation (DERS) | 0.043 (0.014) | [0.016, 0.070] | 0.24 [0.09, 0.39] | .002 | .032 |
|  | Impulsivity (BIS-10) | 0.014 (0.022) | [-0.029, 0.058] | 0.04 [-0.09, 0.18] | .516 | .933 |
|  | Dissociation (DES) | 0.023 (0.021) | [-0.018, 0.065] | 0.07 [-0.05, 0.19] | .271 | .712 |
|  | Childhood maltreatment (CTQ) | 0.029 (0.013) | [0.003, 0.056] | 0.13 [0.01, 0.24] | .032 | .298 |
|  | Current affective disorder | 0.457 (0.556) | [-0.641, 1.554] | 0.10 [-0.15, 0.35] | .413 | .889 |
|  | Lifetime affective disorder | -0.242 (0.656) | [-1.539, 1.054] | -0.06 [-0.35, 0.24] | .712 | .959 |
|  | Lifetime psychotic disorder | -0.046 (1.444) | [-2.893, 2.802] | -0.01 [-0.66, 0.64] | .975 | .991 |
|  | Lifetime substance use disorder | -0.114 (0.592) | [-1.281, 1.053] | -0.03 [-0.29, 0.24] | .848 | .989 |
|  | Current anxiety disorder | 0.202 (0.533) | [-0.851, 1.255] | 0.05 [-0.19, 0.28] | .705 | .959 |
|  | Lifetime anxiety disorder | 0.150 (0.552) | [-0.940, 1.239] | 0.03 [-0.21, 0.28] | .786 | .959 |
|  | Current somatoform disorder | 0.052 (1.211) | [-2.339, 2.442] | 0.01 [-0.53, 0.55] | .966 | .991 |
|  | Current eating disorder | 0.605 (0.589) | [-0.555, 1.765] | 0.14 [-0.13, 0.40] | .305 | .712 |
|  | Lifetime eating disorder | 0.680 (0.505) | [-0.317, 1.677] | 0.15 [-0.07, 0.38] | .180 | .712 |
|  | Lifetime adjustment or other disorder | -0.920 (1.474) | [-3.825, 1.985] | -0.21 [-0.87, 0.45] | .533 | .933 |
| BPD symptom severity (BSL-23) | Age | 0.010 (0.007) | [-0.004, 0.024] | 0.09 [-0.04, 0.21] | .175 | .919 |
|  | Female sex | -0.021 (0.140) | [-0.296, 0.255] | -0.02 [-0.32, 0.27] | .881 | .969 |
|  | Migration background | -0.032 (0.277) | [-0.579, 0.515] | -0.04 [-0.63, 0.56] | .908 | .969 |
|  | Educational attainment | -0.071 (0.073) | [-0.215, 0.073] | -0.06 [-0.17, 0.06] | .331 | .969 |
|  | Married or marriage-like relationship | 0.028 (0.127) | [-0.223, 0.279] | 0.03 [-0.24, 0.30] | .827 | .969 |
|  | Employed or in education | -0.282 (0.188) | [-0.657, 0.093] | -0.30 [-0.71, 0.10] | .137 | .919 |
|  | Independent living | 0.018 (0.137) | [-0.252, 0.289] | 0.02 [-0.27, 0.31] | .893 | .969 |
|  | Number of biological children | 0.066 (0.055) | [-0.042, 0.174] | 0.07 [-0.04, 0.18] | .227 | .919 |
|  | Psychopharmacological treatment | 0.123 (0.136) | [-0.146, 0.392] | 0.13 [-0.16, 0.42] | .368 | .969 |
|  | Inpatient psychiatric treatment | 0.035 (0.112) | [-0.187, 0.256] | 0.04 [-0.20, 0.28] | .758 | .969 |
|  | Outpatient psychotherapy | 0.088 (0.097) | [-0.103, 0.279] | 0.10 [-0.11, 0.30] | .365 | .969 |
|  | Lifetime suicide attempt | 0.018 (0.098) | [-0.176, 0.212] | 0.02 [-0.19, 0.23] | .854 | .969 |
|  | BPD diagnosis (IPDE) | 0.066 (0.167) | [-0.265, 0.396] | 0.07 [-0.29, 0.43] | .695 | .969 |
|  | Number of BPD criteria (IPDE) | 0.052 (0.029) | [-0.005, 0.109] | 0.13 [-0.01, 0.27] | .073 | .919 |
|  | Dimensional BPD score (IPDE) | 0.023 (0.015) | [-0.007, 0.054] | 0.10 [-0.03, 0.24] | .135 | .919 |
|  | BPD symptom severity (ZAN-BPD) | 0.128 (0.109) | [-0.089, 0.345] | 0.09 [-0.06, 0.25] | .245 | .919 |
|  | Emotion dysregulation (DERS) | 0.004 (0.003) | [-0.002, 0.009] | 0.10 [-0.05, 0.25] | .184 | .919 |
|  | Impulsivity (BIS-10) | 0.003 (0.005) | [-0.006, 0.012] | 0.04 [-0.09, 0.17] | .552 | .969 |
|  | Dissociation (DES) | -0.001 (0.005) | [-0.012, 0.009] | -0.02 [-0.16, 0.13] | .822 | .969 |
|  | Childhood maltreatment (CTQ) | 0.007 (0.003) | [0.001, 0.014] | 0.15 [0.02, 0.29] | .028 | .849 |
|  | Current affective disorder | -0.017 (0.127) | [-0.269, 0.236] | -0.02 [-0.29, 0.25] | .896 | .969 |
|  | Lifetime affective disorder | -0.028 (0.131) | [-0.286, 0.230] | -0.03 [-0.31, 0.25] | .831 | .969 |
|  | Lifetime psychotic disorder | 0.010 (0.270) | [-0.522, 0.542] | 0.01 [-0.56, 0.59] | .969 | .969 |
|  | Lifetime substance use disorder | 0.057 (0.114) | [-0.167, 0.282] | 0.06 [-0.18, 0.30] | .615 | .969 |
|  | Current anxiety disorder | -0.025 (0.126) | [-0.275, 0.224] | -0.03 [-0.30, 0.24] | .841 | .969 |
|  | Lifetime anxiety disorder | 0.009 (0.113) | [-0.215, 0.234] | 0.01 [-0.23, 0.25] | .934 | .969 |
|  | Current somatoform disorder | -0.181 (0.232) | [-0.639, 0.277] | -0.20 [-0.69, 0.30] | .436 | .969 |
|  | Current eating disorder | 0.009 (0.124) | [-0.237, 0.254] | 0.01 [-0.26, 0.27] | .944 | .969 |
|  | Lifetime eating disorder | 0.063 (0.100) | [-0.133, 0.260] | 0.07 [-0.14, 0.28] | .526 | .969 |
|  | Lifetime adjustment or other disorder | -0.152 (0.280) | [-0.703, 0.400] | -0.16 [-0.76, 0.43] | .588 | .969 |
| BPD symptom severity (ZAN-BPD) | Age | 0.012 (0.006) | [0.000, 0.024] | 0.14 [0.00, 0.28] | .048 | .330 |
|  | Female sex | -0.140 (0.130) | [-0.397, 0.117] | -0.20 [-0.56, 0.16] | .283 | .707 |
|  | Migration background | 0.032 (0.252) | [-0.466, 0.530] | 0.04 [-0.66, 0.74] | .900 | .907 |
|  | Educational attainment | -0.096 (0.077) | [-0.249, 0.057] | -0.10 [-0.25, 0.06] | .217 | .652 |
|  | Married or marriage-like relationship | 0.085 (0.113) | [-0.138, 0.309] | 0.12 [-0.20, 0.44] | .452 | .745 |
|  | Employed or in education | -0.273 (0.191) | [-0.658, 0.111] | -0.38 [-0.92, 0.15] | .160 | .598 |
|  | Independent living | -0.033 (0.113) | [-0.257, 0.191] | -0.05 [-0.36, 0.27] | .773 | .865 |
|  | Number of biological children | 0.083 (0.054) | [-0.024, 0.190] | 0.11 [-0.03, 0.25] | .126 | .540 |
|  | Psychopharmacological treatment | 0.028 (0.099) | [-0.167, 0.223] | 0.04 [-0.24, 0.31] | .778 | .865 |
|  | Inpatient psychiatric treatment | -0.057 (0.094) | [-0.241, 0.128] | -0.08 [-0.34, 0.18] | .547 | .782 |
|  | Outpatient psychotherapy | 0.079 (0.084) | [-0.087, 0.245] | 0.11 [-0.12, 0.35] | .349 | .745 |
|  | Lifetime suicide attempt | 0.015 (0.084) | [-0.150, 0.181] | 0.02 [-0.21, 0.25] | .854 | .907 |
|  | BPD diagnosis (IPDE) | 0.103 (0.145) | [-0.182, 0.388] | 0.14 [-0.26, 0.54] | .477 | .745 |
|  | Number of BPD criteria (IPDE) | 0.060 (0.028) | [0.005, 0.114] | 0.19 [0.02, 0.37] | .033 | .330 |
|  | Dimensional BPD score (IPDE) | 0.031 (0.014) | [0.003, 0.060] | 0.18 [0.02, 0.35] | .033 | .330 |
|  | BPD symptom severity (BSL-23) | 0.229 (0.068) | [0.094, 0.364] | 0.29 [0.12, 0.46] | .001 | .032 |
|  | Emotion dysregulation (DERS) | 0.005 (0.002) | [0.000, 0.010] | 0.17 [-0.00, 0.34] | .055 | .330 |
|  | Impulsivity (BIS-10) | 0.003 (0.003) | [-0.004, 0.009] | 0.05 [-0.08, 0.18] | .436 | .745 |
|  | Dissociation (DES) | 0.002 (0.004) | [-0.006, 0.009] | 0.03 [-0.10, 0.15] | .659 | .865 |
|  | Childhood maltreatment (CTQ) | 0.004 (0.002) | [0.000, 0.008] | 0.11 [-0.01, 0.23] | .078 | .392 |
|  | Current affective disorder | -0.029 (0.097) | [-0.220, 0.163] | -0.04 [-0.31, 0.23] | .768 | .865 |
|  | Lifetime affective disorder | 0.029 (0.102) | [-0.172, 0.230] | 0.04 [-0.24, 0.32] | .775 | .865 |
|  | Lifetime psychotic disorder | -0.027 (0.233) | [-0.486, 0.432] | -0.04 [-0.68, 0.61] | .907 | .907 |
|  | Lifetime substance use disorder | 0.078 (0.099) | [-0.117, 0.274] | 0.11 [-0.16, 0.38] | .431 | .745 |
|  | Current anxiety disorder | 0.108 (0.084) | [-0.059, 0.274] | 0.15 [-0.08, 0.39] | .203 | .652 |
|  | Lifetime anxiety disorder | 0.059 (0.086) | [-0.111, 0.229] | 0.08 [-0.16, 0.32] | .497 | .745 |
|  | Current somatoform disorder | -0.141 (0.197) | [-0.529, 0.246] | -0.20 [-0.74, 0.35] | .473 | .745 |
|  | Current eating disorder | 0.072 (0.101) | [-0.127, 0.272] | 0.10 [-0.18, 0.38] | .477 | .745 |
|  | Lifetime eating disorder | 0.089 (0.082) | [-0.073, 0.250] | 0.12 [-0.10, 0.35] | .282 | .707 |
|  | Lifetime adjustment or other disorder | -0.078 (0.240) | [-0.551, 0.395] | -0.11 [-0.78, 0.55] | .746 | .865 |
| Married or marriage-like relationship | Age | -0.023 (0.025) | [-0.072, 0.027] | 0.83 [0.55, 1.24] | .362 | .901 |
|  | Female sex | 0.138 (0.485) | [-0.820, 1.096] | 1.15 [0.44, 2.99] | .776 | .901 |
|  | Migration background | 0.702 (0.769) | [-0.816, 2.219] | 2.02 [0.44, 9.20] | .363 | .901 |
|  | Educational attainment | -0.134 (0.205) | [-0.538, 0.270] | 0.91 [0.68, 1.21] | .514 | .901 |
|  | Employed or in education | 0.320 (0.565) | [-0.807, 1.447] | 1.38 [0.45, 4.25] | .573 | .901 |
|  | Independent living | -0.185 (0.421) | [-1.017, 0.647] | 0.83 [0.36, 1.91] | .661 | .901 |
|  | Number of biological children | -0.033 (0.200) | [-0.428, 0.363] | 0.97 [0.67, 1.41] | .871 | .901 |
|  | Psychopharmacological treatment | -0.382 (0.375) | [-1.122, 0.358] | 0.68 [0.33, 1.43] | .310 | .901 |
|  | Inpatient psychiatric treatment | -0.112 (0.335) | [-0.774, 0.549] | 0.89 [0.46, 1.73] | .738 | .901 |
|  | Outpatient psychotherapy | -0.080 (0.299) | [-0.670, 0.511] | 0.92 [0.51, 1.67] | .791 | .901 |
|  | Lifetime suicide attempt | 0.222 (0.308) | [-0.386, 0.830] | 1.25 [0.68, 2.29] | .472 | .901 |
|  | BPD diagnosis (IPDE) | -0.530 (0.434) | [-1.388, 0.329] | 0.59 [0.25, 1.39] | .224 | .901 |
|  | Number of BPD criteria (IPDE) | -0.096 (0.071) | [-0.236, 0.044] | 0.80 [0.58, 1.11] | .176 | .901 |
|  | Dimensional BPD score (IPDE) | -0.050 (0.038) | [-0.124, 0.025] | 0.81 [0.60, 1.11] | .189 | .901 |
|  | BPD symptom severity (BSL-23) | -0.258 (0.180) | [-0.614, 0.098] | 0.79 [0.58, 1.09] | .155 | .901 |
|  | BPD symptom severity (ZAN-BPD) | -0.294 (0.250) | [-0.789, 0.201] | 0.82 [0.59, 1.14] | .242 | .901 |
|  | Emotion dysregulation (DERS) | -0.020 (0.008) | [-0.035, -0.005] | 0.62 [0.43, 0.89] | .010 | .310 |
|  | Impulsivity (BIS-10) | -0.011 (0.011) | [-0.033, 0.010] | 0.86 [0.64, 1.14] | .294 | .901 |
|  | Dissociation (DES) | -0.008 (0.012) | [-0.031, 0.016] | 0.91 [0.67, 1.22] | .509 | .901 |
|  | Childhood maltreatment (CTQ) | -0.001 (0.008) | [-0.017, 0.014] | 0.97 [0.72, 1.31] | .850 | .901 |
|  | Current affective disorder | -0.266 (0.321) | [-0.898, 0.367] | 0.77 [0.41, 1.44] | .408 | .901 |
|  | Lifetime affective disorder | 0.139 (0.360) | [-0.570, 0.849] | 1.15 [0.56, 2.34] | .699 | .901 |
|  | Lifetime psychotic disorder | -0.298 (0.968) | [-2.206, 1.610] | 0.74 [0.11, 5.00] | .759 | .901 |
|  | Lifetime substance use disorder | 0.122 (0.334) | [-0.537, 0.781] | 1.13 [0.58, 2.18] | .715 | .901 |
|  | Current anxiety disorder | -0.129 (0.302) | [-0.724, 0.467] | 0.88 [0.48, 1.59] | .670 | .901 |
|  | Lifetime anxiety disorder | -0.150 (0.324) | [-0.790, 0.490] | 0.86 [0.45, 1.63] | .644 | .901 |
|  | Current somatoform disorder | -0.246 (0.741) | [-1.708, 1.216] | 0.78 [0.18, 3.37] | .740 | .901 |
|  | Current eating disorder | 0.065 (0.334) | [-0.592, 0.723] | 1.07 [0.55, 2.06] | .845 | .901 |
|  | Lifetime eating disorder | 0.061 (0.281) | [-0.492, 0.615] | 1.06 [0.61, 1.85] | .828 | .901 |
|  | Lifetime adjustment or other disorder | 0.069 (0.839) | [-1.583, 1.722] | 1.07 [0.20, 5.60] | .934 | .934 |
| Employed or in education | Age | -0.076 (0.022) | [-0.120, -0.032] | 0.54 [0.38, 0.77] | <.001 | .026 |
|  | Female sex | 0.072 (0.429) | [-0.776, 0.919] | 1.07 [0.46, 2.51] | .868 | .953 |
|  | Migration background | 0.047 (0.847) | [-1.625, 1.720] | 1.05 [0.20, 5.58] | .956 | .965 |
|  | Educational attainment | 0.179 (0.215) | [-0.247, 0.605] | 1.14 [0.84, 1.54] | .407 | .643 |
|  | Married or marriage-like relationship | -0.136 (0.360) | [-0.848, 0.575] | 0.87 [0.43, 1.78] | .706 | .847 |
|  | Independent living | -0.395 (0.401) | [-1.187, 0.397] | 0.67 [0.30, 1.49] | .327 | .613 |
|  | Number of biological children | -0.240 (0.174) | [-0.585, 0.106] | 0.80 [0.58, 1.10] | .172 | .506 |
|  | Psychopharmacological treatment | -0.296 (0.341) | [-0.967, 0.376] | 0.74 [0.38, 1.46] | .386 | .643 |
|  | Inpatient psychiatric treatment | -0.315 (0.311) | [-0.929, 0.298] | 0.73 [0.40, 1.35] | .312 | .613 |
|  | Outpatient psychotherapy | 0.043 (0.309) | [-0.566, 0.652] | 1.04 [0.57, 1.92] | .889 | .953 |
|  | Lifetime suicide attempt | 0.153 (0.286) | [-0.412, 0.717] | 1.17 [0.66, 2.05] | .594 | .743 |
|  | BPD diagnosis (IPDE) | -0.635 (0.495) | [-1.614, 0.345] | 0.53 [0.20, 1.41] | .202 | .506 |
|  | Number of BPD criteria (IPDE) | -0.162 (0.080) | [-0.320, -0.004] | 0.69 [0.48, 0.99] | .044 | .293 |
|  | Dimensional BPD score (IPDE) | -0.090 (0.045) | [-0.179, 0.000] | 0.69 [0.47, 1.00] | .049 | .293 |
|  | BPD symptom severity (BSL-23) | -0.617 (0.199) | [-1.010, -0.223] | 0.57 [0.40, 0.82] | .002 | .036 |
|  | BPD symptom severity (ZAN-BPD) | -0.382 (0.233) | [-0.842, 0.078] | 0.78 [0.57, 1.05] | .103 | .344 |
|  | Emotion dysregulation (DERS) | -0.013 (0.006) | [-0.026, 0.000] | 0.73 [0.53, 0.99] | .044 | .293 |
|  | Impulsivity (BIS-10) | -0.013 (0.011) | [-0.034, 0.008] | 0.84 [0.63, 1.11] | .223 | .516 |
|  | Dissociation (DES) | -0.010 (0.011) | [-0.032, 0.011] | 0.88 [0.67, 1.16] | .350 | .618 |
|  | Childhood maltreatment (CTQ) | -0.008 (0.007) | [-0.023, 0.007] | 0.85 [0.64, 1.14] | .278 | .595 |
|  | Current affective disorder | -0.539 (0.298) | [-1.126, 0.047] | 0.58 [0.32, 1.05] | .071 | .306 |
|  | Lifetime affective disorder | -0.489 (0.379) | [-1.235, 0.258] | 0.61 [0.29, 1.29] | .199 | .506 |
|  | Lifetime psychotic disorder | 0.516 (0.939) | [-1.335, 2.367] | 1.68 [0.26, 10.66] | .583 | .743 |
|  | Lifetime substance use disorder | 0.091 (0.342) | [-0.584, 0.766] | 1.09 [0.56, 2.15] | .791 | .912 |
|  | Current anxiety disorder | -0.540 (0.292) | [-1.116, 0.037] | 0.58 [0.33, 1.04] | .066 | .306 |
|  | Lifetime anxiety disorder | -0.535 (0.318) | [-1.162, 0.092] | 0.59 [0.31, 1.10] | .094 | .344 |
|  | Current somatoform disorder | 0.029 (0.658) | [-1.267, 1.325] | 1.03 [0.28, 3.76] | .965 | .965 |
|  | Current eating disorder | -0.250 (0.338) | [-0.916, 0.416] | 0.78 [0.40, 1.52] | .460 | .657 |
|  | Lifetime eating disorder | -0.201 (0.287) | [-0.768, 0.365] | 0.82 [0.46, 1.44] | .484 | .660 |
|  | Lifetime adjustment or other disorder | -0.601 (0.758) | [-2.094, 0.893] | 0.55 [0.12, 2.44] | .429 | .644 |
| Psychopharmacological treatment | Age | 0.002 (0.019) | [-0.035, 0.039] | 1.02 [0.75, 1.38] | .902 | .977 |
|  | Female sex | -0.041 (0.401) | [-0.834, 0.751] | 0.96 [0.43, 2.12] | .918 | .977 |
|  | Migration background | -0.353 (0.813) | [-1.959, 1.252] | 0.70 [0.14, 3.50] | .665 | .977 |
|  | Educational attainment | -0.111 (0.188) | [-0.483, 0.260] | 0.92 [0.71, 1.20] | .555 | .977 |
|  | Married or marriage-like relationship | -0.100 (0.375) | [-0.843, 0.642] | 0.90 [0.43, 1.90] | .789 | .977 |
|  | Employed or in education | -0.251 (0.471) | [-1.191, 0.689] | 0.78 [0.30, 1.99] | .596 | .977 |
|  | Independent living | -0.301 (0.362) | [-1.015, 0.413] | 0.74 [0.36, 1.51] | .406 | .977 |
|  | Number of biological children | 0.033 (0.163) | [-0.290, 0.357] | 1.03 [0.76, 1.40] | .838 | .977 |
|  | Inpatient psychiatric treatment | 0.090 (0.292) | [-0.487, 0.666] | 1.09 [0.61, 1.95] | .760 | .977 |
|  | Outpatient psychotherapy | 0.050 (0.267) | [-0.476, 0.577] | 1.05 [0.62, 1.78] | .851 | .977 |
|  | Lifetime suicide attempt | -0.184 (0.257) | [-0.690, 0.323] | 0.83 [0.50, 1.38] | .476 | .977 |
|  | BPD diagnosis (IPDE) | -0.155 (0.387) | [-0.920, 0.610] | 0.86 [0.40, 1.84] | .690 | .977 |
|  | Number of BPD criteria (IPDE) | -0.016 (0.061) | [-0.137, 0.104] | 0.96 [0.73, 1.27] | .791 | .977 |
|  | Dimensional BPD score (IPDE) | -0.007 (0.034) | [-0.073, 0.060] | 0.97 [0.74, 1.28] | .847 | .977 |
|  | BPD symptom severity (BSL-23) | 0.005 (0.153) | [-0.298, 0.307] | 1.00 [0.77, 1.32] | .975 | .977 |
|  | BPD symptom severity (ZAN-BPD) | -0.178 (0.210) | [-0.593, 0.237] | 0.89 [0.67, 1.17] | .399 | .977 |
|  | Emotion dysregulation (DERS) | -0.002 (0.006) | [-0.013, 0.009] | 0.95 [0.72, 1.26] | .714 | .977 |
|  | Impulsivity (BIS-10) | 0.002 (0.010) | [-0.018, 0.023] | 1.03 [0.79, 1.36] | .821 | .977 |
|  | Dissociation (DES) | 0.003 (0.010) | [-0.017, 0.023] | 1.04 [0.81, 1.34] | .748 | .977 |
|  | Childhood maltreatment (CTQ) | 0.002 (0.007) | [-0.012, 0.017] | 1.04 [0.79, 1.38] | .760 | .977 |
|  | Current affective disorder | -0.009 (0.295) | [-0.590, 0.573] | 0.99 [0.55, 1.77] | .977 | .977 |
|  | Lifetime affective disorder | 0.025 (0.322) | [-0.609, 0.659] | 1.03 [0.54, 1.93] | .937 | .977 |
|  | Lifetime psychotic disorder | 1.038 (0.767) | [-0.473, 2.549] | 2.82 [0.62, 12.80] | .177 | .977 |
|  | Lifetime substance use disorder | 0.142 (0.298) | [-0.445, 0.730] | 1.15 [0.64, 2.08] | .633 | .977 |
|  | Current anxiety disorder | 0.015 (0.267) | [-0.511, 0.541] | 1.01 [0.60, 1.72] | .955 | .977 |
|  | Lifetime anxiety disorder | 0.031 (0.292) | [-0.545, 0.607] | 1.03 [0.58, 1.83] | .916 | .977 |
|  | Current somatoform disorder | 0.064 (0.603) | [-1.124, 1.252] | 1.07 [0.33, 3.50] | .916 | .977 |
|  | Current eating disorder | 0.050 (0.323) | [-0.587, 0.686] | 1.05 [0.56, 1.99] | .877 | .977 |
|  | Lifetime eating disorder | -0.020 (0.250) | [-0.512, 0.472] | 0.98 [0.60, 1.60] | .936 | .977 |
|  | Lifetime adjustment or other disorder | 0.202 (0.767) | [-1.310, 1.714] | 1.22 [0.27, 5.55] | .793 | .977 |
| Outpatient psychotherapy | Age | 0.038 (0.020) | [-0.002, 0.077] | 1.36 [0.98, 1.88] | .062 | .963 |
|  | Female sex | -0.106 (0.385) | [-0.866, 0.653] | 0.90 [0.42, 1.92] | .783 | .963 |
|  | Migration background | -0.490 (0.856) | [-2.182, 1.201] | 0.61 [0.11, 3.32] | .568 | .963 |
|  | Educational attainment | -0.009 (0.190) | [-0.383, 0.365] | 0.99 [0.76, 1.30] | .963 | .963 |
|  | Married or marriage-like relationship | 0.243 (0.364) | [-0.478, 0.963] | 1.27 [0.62, 2.62] | .506 | .963 |
|  | Employed or in education | -0.330 (0.455) | [-1.238, 0.577] | 0.72 [0.29, 1.78] | .471 | .963 |
|  | Independent living | 0.189 (0.360) | [-0.523, 0.900] | 1.21 [0.59, 2.46] | .601 | .963 |
|  | Number of biological children | 0.164 (0.150) | [-0.133, 0.461] | 1.17 [0.88, 1.54] | .277 | .963 |
|  | Psychopharmacological treatment | 0.335 (0.314) | [-0.285, 0.955] | 1.40 [0.75, 2.60] | .288 | .963 |
|  | Inpatient psychiatric treatment | 0.079 (0.289) | [-0.491, 0.649] | 1.08 [0.61, 1.91] | .785 | .963 |
|  | Lifetime suicide attempt | -0.068 (0.271) | [-0.602, 0.467] | 0.94 [0.55, 1.59] | .803 | .963 |
|  | BPD diagnosis (IPDE) | 0.398 (0.389) | [-0.370, 1.165] | 1.49 [0.69, 3.21] | .308 | .963 |
|  | Number of BPD criteria (IPDE) | 0.078 (0.061) | [-0.042, 0.198] | 1.20 [0.91, 1.58] | .203 | .963 |
|  | Dimensional BPD score (IPDE) | 0.038 (0.034) | [-0.029, 0.106] | 1.17 [0.89, 1.55] | .265 | .963 |
|  | BPD symptom severity (BSL-23) | 0.196 (0.155) | [-0.111, 0.503] | 1.19 [0.91, 1.57] | .209 | .963 |
|  | BPD symptom severity (ZAN-BPD) | 0.410 (0.230) | [-0.045, 0.864] | 1.31 [0.97, 1.78] | .077 | .963 |
|  | Emotion dysregulation (DERS) | 0.006 (0.006) | [-0.006, 0.018] | 1.16 [0.87, 1.55] | .314 | .963 |
|  | Impulsivity (BIS-10) | 0.001 (0.010) | [-0.018, 0.020] | 1.02 [0.79, 1.32] | .894 | .963 |
|  | Dissociation (DES) | 0.004 (0.010) | [-0.016, 0.024] | 1.06 [0.82, 1.36] | .667 | .963 |
|  | Childhood maltreatment (CTQ) | 0.005 (0.007) | [-0.008, 0.019] | 1.11 [0.85, 1.45] | .438 | .963 |
|  | Current affective disorder | 0.085 (0.270) | [-0.446, 0.617] | 1.09 [0.64, 1.85] | .752 | .963 |
|  | Lifetime affective disorder | 0.205 (0.326) | [-0.439, 0.849] | 1.23 [0.65, 2.34] | .530 | .963 |
|  | Lifetime psychotic disorder | -0.404 (0.813) | [-2.008, 1.200] | 0.67 [0.13, 3.32] | .620 | .963 |
|  | Lifetime substance use disorder | -0.220 (0.289) | [-0.789, 0.350] | 0.80 [0.45, 1.42] | .448 | .963 |
|  | Current anxiety disorder | -0.038 (0.273) | [-0.578, 0.502] | 0.96 [0.56, 1.65] | .890 | .963 |
|  | Lifetime anxiety disorder | 0.084 (0.279) | [-0.467, 0.635] | 1.09 [0.63, 1.89] | .763 | .963 |
|  | Current somatoform disorder | -0.029 (0.610) | [-1.233, 1.174] | 0.97 [0.29, 3.23] | .962 | .963 |
|  | Current eating disorder | 0.024 (0.297) | [-0.561, 0.608] | 1.02 [0.57, 1.84] | .936 | .963 |
|  | Lifetime eating disorder | 0.158 (0.248) | [-0.332, 0.648] | 1.17 [0.72, 1.91] | .525 | .963 |
|  | Lifetime adjustment or other disorder | -0.761 (0.971) | [-2.678, 1.155] | 0.47 [0.07, 3.17] | .434 | .963 |

*Note.* Estimates were pooled across 100 multiply imputed data sets (*N* = 410). *b* = unstandardized coefficient (linear regression for continuous outcomes, logistic regression for binary outcomes); β = standardized coefficient (outcome and continuous predictors *z*-standardized, binary predictors coded 0/1); OR_SD_ = odds ratio per standard deviation of the predictor; *p*_FDR_ = Benjamini–Hochberg-adjusted *p* value within outcome. For IPDE diagnosis outcomes, the dimensional baseline score served as the autoregressor. BIS-10, Barratt Impulsiveness Scale, Version 10; BPD, borderline personality disorder; BSL-23, Borderline Symptom List, short version; CI, confidence interval; CTQ, Childhood Trauma Questionnaire; DERS, Difficulties in Emotion Regulation Scale; DES, Dissociative Experiences Scale; FDR, false discovery rate; IPDE, International Personality Disorder Examination; SE, standard error; ZAN-BPD, Zanarini Rating Scale for Borderline Personality Disorder.

^a^ Estimate not interpretable due to quasi-complete separation (rare predictor category); coefficient, CI, and OR omitted.

**Table S7.** *Internal Consistency of Self-Report Measures and Clinician Ratings*

| Scale | Occasion | *k* | α | Min. *r*_it_ | *n* |
| --- | --- | --- | --- | --- | --- |
| ZAN-BPD | Baseline | 9 | .78 | .35 | 400 |
| ZAN-BPD | Follow-up | 9 | .84 | .40 | 185 |
| BSL-23 | Baseline | 23 | .95 | .26 | 353 |
| BSL-23 | Follow-up | 23 | .96 | .32 | 183 |
| DERS | Baseline | 36 | .93 | .02 | 360 |
| DES | Baseline | 44 | .94 | .30 | 362 |
| BIS-10 | Baseline | 34 | .85 | -.01 | 362 |
| CTQ | Baseline | 25 | .93 | .32 | 335 |

*Note.* α = Cronbach's alpha; *k* = number of items; Min. *r*_it_ = lowest corrected item–total correlation; *n* = participants with complete item data. Baseline = T0; Follow-up = T1. Follow-up internal consistencies are reported only for instruments whose follow-up data entered the analyses (BSL-23, ZAN-BPD). CTQ total α was computed across all 25 items with reverse-keyed items recoded. BIS-10, Barratt Impulsiveness Scale, Version 10; BSL-23, Borderline Symptom List, short version; CTQ, Childhood Trauma Questionnaire; DERS, Difficulties in Emotion Regulation Scale; DES, Dissociative Experiences Scale; ZAN-BPD, Zanarini Rating Scale for Borderline Personality Disorder.

5.	World Health Organization. International Statistical Classification of Diseases and Related Health Problems, Tenth Revision. 2nd ed. Geneva: World Health Organization; 2004.

6.	American Psychiatric Association. Diagnostic and Statistical Manual Of Mental Disorders 4ed. Washington, DC: American Psychiatric Association; 1994.

15.	Wittchen H-U, Zaudig M, Fydrich T. SKID: Strukturiertes Klinisches Interview für DSM-IV. Achse I und II. Göttingen: Hogrefe; 1997.

16.	van Buuren S, Groothuis-Oudshoorn K. mice: Multivariate Imputation by Chained Equations in R. Journal of Statistical Software. 2011;45(3):1 – 67.

17.	Collins LM, Schafer JL, Kam CM. A comparison of inclusive and restrictive strategies in modern missing data procedures. Psychol Methods. 2001;6(4):330–51.

18.	van Buuren S. Flexible imputation of missing data. 2nd ed. Boca Raton, FL: CRC Press; 2018.

19.	Rubin DB. Multiple imputation for nonresponse in surveys. New York, NY: Wiley; 1987.

20.	Lakens D. Calculating and reporting effect sizes to facilitate cumulative science: a practical primer for t-tests and ANOVAs. Front Psychol. 2013;4:863.

21.	Benjamini Y, Hochberg Y. Controlling the False Discovery Rate: A Practical and Powerful Approach to Multiple Testing. Journal of the Royal Statistical Society: Series B (Methodological). 1995;57(1):289–300.

22.	Graham JW. Adding Missing-Data-Relevant Variables to FIML-Based Structural Equation Models. Structural Equation Modeling: A Multidisciplinary Journal. 2003;10(1):80–100.

23.	Rosseel Y. lavaan: An R Package for Structural Equation Modeling. Journal of Statistical Software. 2012;48(2):1 – 36.
